# Step change in glaucoma polygenic risk score performance enables clinical utility and disease prediction across all major ancestries

**DOI:** 10.64898/2026.01.23.26344675

**Authors:** Stuart MacGregor, Guiyan Ni, Mathias Seviiri, Antonia Kolovos, Ngoc Quynh Le, Mark M Hassall, Josh Schmidt, Puya Gharahkhani, Victoria Tang, Emmanuelle Souzeau, Ayub Qassim, Priyanka Nandakumar, Suyash Shringarpure, Julie M Granka, Jessica N Cooke Bailey, Tin Aung, John Landers, Stuart L Graham, Paul R Healey, Anthony P Khawaja, Louis R Pasquale, Janey L Wiggs, Joan O’Brien, Robert N Weinreb, Colin E Willoughby, Andrew Lotery, Paul Mitchell, David A Mackey, Alex W Hewitt, Nick Haan, Owen M Siggs, Jamie E Craig

**Affiliations:** Statistical Genetics, QIMR Berghofer, 300 Herston Road, Herston, QLD, 4006, Australia; Faculty of Medicine, University of Queensland, 288 Herston Road, Herston, QLD, 4006, Australia; Seonix Bio, North Terrace, Adelaide, SA, 5000, Australia; Department of Ophthalmology, Flinders Medical Centre, Flinders University, Sturt Rd, Adelaide, SA, 5042, Australia; Department of Ophthalmology, Flinders Medical Centre, Flinders University, Sturt Rd, Bedford Park, SA, 5042, Australia; QIMR Berghofer, 300 Herston Road, Herston, QLD, 4006, Australia; University of Queensland, 288 Herston Road, Herston, QLD, 4006, Australia; Department of Ophthalmology, Flinders Medical Centre, Flinders Health and Medical Research Institute, Sturt Rd, Bedford Park, SA, 5042, Australia; 23andMe, Inc., 223 N Mathilda Ave, Sunnyvale, CA, 94086, USA; Department of Pharmacology & Toxicology, Center for Health Disparities, Brody School of Medicine, East Carolina University, 600 Moye Blvd, Greenville, NC, 27834, USA; Singapore Eye Research Institute and Singapore National Eye Center, 20 College Rd, Level 6 Discovery Tower, Singapore, 169856, Singapore; Duke-NUS Medical School, 8 College Rd, Singapore, 169856, Singapore; Yong Loo Lin School of Medicine, National University of Singapore, 10 Medical Dr, Singapore, 117597, Singapore; Ophthalmology and Vision Science, Faculty of Medicine, Health and Human Sciences, Macquarie University, L4/Suite 401 2 Technology Pl Macquarie University, Sydney, NSW, 2109, Australia; Clinical Ophthalmology, Westmead Clinical School, University of Sydney, Parramatta and City Roads, Sydney, NSW, 2006, Australia; Moorfields Eye Hospital NHS Foundation Trust and UCL Institute of Ophthalmology, NIHR Biomedical Research Centre, 11-43 Bath Street, London, EC1V 9EL, UK; New York Eye and Ear Infirmary of Mount Sinai, Icahn School of Medicine at Mount Sinai, 1 Gustave L. Levy Place, New York, NY, 10029-5674, USA; Department of Ophthalmology, Massachusetts Eye and Ear, Harvard Medical School, Boston, MA, USA; Broad Institute of Harvard and MIT, Cambridge, MA, USA; Department of Ophthalmology, University of Pennsylvania, Philadelphia, PA, 19104, USA; Viterbi Family Department of Ophthalmology, Hamilton Glaucoma Center, Shiley Eye Institute, University of California San Diego, 9415 Campus Point Drive, La Jolla, California, 92093-0946, USA; Biomedical Sciences Research Institute, Centre for Genomic Medicine, Ulster University, Coleraine, Northern Ireland, UK; University Hospital Southampton NHS Foundation Trust, Tremona Road, Southampton, SO16 6YD, UK; Faculty of Medicine, University of Southampton, University Road, Southampton, SO17 1BJ, UK; Department of Ophthalmology and Westmead Institute for Medical Research, University of Sydney, Parramatta and City Roads, Sydney, NSW, 2006, Australia; Centre for Ophthalmology and Visual Science, Lions Eye Institute, University of Western Australia, Perth, WA, Australia; Menzies Institute for Medical Research, University of Tasmania, 17 Liverpool Street, Hobart, TAS, 7000, Australia; Centre for Eye Research Australia, University of Melbourne, 200 Victoria Parade, Melbourne, VIC, 3002, Australia; Genomics and Inherited Disease Program, Garvan Institute of Medical Research, 384 Victoria St, Darlinghurst, NSW, 2010, Australia; Faculty of Medicine and Health, University of New South Wales, Sydney, NSW, 2052, Australia

**Author notes:** **Correspondence:** Stuart MacGregor, Statistical Genetics, QIMR Berghofer Medical Research Institute, Brisbane, Australia., 300 Herston Road, Herston, Brisbane, Queensland 4006, Australia. Funding Australian NHMRC, National Institutes of Health.

**Keywords:** glaucoma, polygenic risk score, prediction, multi-ancestry

## Abstract

Glaucoma is the leading cause of irreversible blindness; vision loss is preventable with timely treatment, but early detection is challenging, leaving ∼50% undiagnosed, highlighting the need for improved risk assessment tools. We developed a polygenic risk score (PRS) using data from >6 million individuals. PRS performance was exceptional in European ancestries; top 10% PRS individuals had 10-fold increased risk (OR=10.0) relative to the remainder. Performance remained good across all major ancestry groups; high-PRS individuals were at high absolute risk, especially Africans. High-risk individuals (top 10% PRS) developed glaucoma up to 25 years earlier than those in the lowest 10% and were at 100 times the risk. The PRS also predicted need for treatment escalation in early glaucoma and both prevalent and incident surgery. Risk profiling with this PRS which is clinically available, enables earlier identification and more timely treatment of high-risk individuals for preventable vision loss, with a reduced screening and monitoring burden for those at low-risk.

**Research in Context:** *Evidence before this study:* Glaucoma is the leading cause of irreversible blindness worldwide. Early detection is essential because current treatments cannot restore lost vision. Population-wide screening using conventional risk factors for glaucoma (such as elevated intraocular pressure) is not currently cost-effective, though a better stratification method could improve early diagnosis rates and reduce over-monitoring of lower risk cases. Glaucoma is one of the most heritable common complex diseases, suggesting that genetics-based approaches could revolutionize risk stratification. Previous polygenic risk scores (PRS) indexing glaucoma genetic risk were limited by restricted applicability outside European ancestry populations, small discovery sample sizes, modest predictive power, poor clinical availability. The inability to cover a broad range of ancestries limited the practical clinical application of earlier versions of glaucoma PRS for risk stratification and patient management.

*Added value of this study:* A novel PRS, trained on >6 million individuals, has significantly improved glaucoma risk prediction across major ancestry groups, identifying high-risk individuals (top 10% of risk versus remainder) with odds ratios up to 10 in Europeans and 3.4–7.5 in non-Europeans (larger than for any other common complex disease). The excellent PRS risk stratification remained after adjusting for intraocular pressure; adding PRS to a prediction model with age, sex and intraocular pressure increased the Area Under the Curve for predicting glaucoma status from 0.63 to 0.82. High-risk individuals (top 10% PRS) developed glaucoma up to 25 years earlier than those in the lowest 10% and were at 100 times greater risk. The PRS also predicted structural progression, the need for treatment initiation and escalation in early glaucoma, both prevalent and incident incisional surgery for glaucoma, and the likelihood of glaucoma being present in first-degree relatives. These improvements impart strong clinical utility for population risk stratification, and the personalised management of individuals with clinical features suggestive of early stage glaucoma.

*Implications of all the available evidence:* This clinically available, enhanced PRS enables accurate identification of both high- and low-risk individuals across diverse populations, supporting earlier diagnosis, targeted monitoring, and timely treatment. It provides a clinical tool which can be applied for precision prevention and management of glaucoma, enabling cost-effective, equitable, genetically-informed glaucoma risk stratification across the entire population.

## INTRODUCTION

Glaucoma manifests through progressive optic nerve degeneration, and is the leading cause of irreversible blindness worldwide^1–3^. Open-angle glaucoma (OAG) is the most common glaucoma subtype^1^, which, in the initial stages, is asymptomatic, with at least half of all cases in the population undiagnosed even in developed countries. The lack of a timely diagnosis can result in permanent loss of vision^4,5^. Early detection is key, as current treatments cannot restore lost vision, with late presentation a major risk factor for blindness^6^. The conventional risk factors are family history, age, and intraocular pressure (IOP)^7^ but these poorly capture glaucoma risk. IOP measurements, while commonly used in ad hoc screening, are unsuitable for screening due to low sensitivity^8^, with 30-80% of those with glaucoma having IOP in the normal range^9–11^. Meticulously collected family history predicts risk well^12^, but oftentimes is poorly annotated and prediction performance is limited in real world studies such as the Blue Mountains Eye Study (Family history area under the curve 0.54^13^). Population-wide screening using conventional risk factors for OAG is not currently cost-effective^7^, though a better stratification method could improve early diagnosis rates and reduce over-monitoring of lower risk cases.

In recent years, as our understanding of the genetics of many complex diseases has increased, polygenic risk scores (PRS) have been developed which can provide an estimate of a person’s inherited risk of disease. Such personalized risk estimates have been developed for a range of complex diseases, with clinical grade PRSs offering moderate risk stratification now being returned to participants (OR per SD between 1.5 and 2.2)^14–16^. Capitalizing on the fact that OAG is one of the most heritable complex human diseases^17–19^, early studies successfully mapped many susceptibility genes^13,20,21^, and developed PRSs for OAG risk and severity^13,4,5^.

While PRSs for OAG (glaucoma henceforth) show particular promise for broader clinical use, key limitations remain. Firstly, existing PRSs only harness a minority of the total genetic contribution to glaucoma^22^. Limited genome-wide association studies (GWAS) sample sizes have been the major factor impeding PRS accuracy^22^. Secondly, due to the limited availability of non-European GWASs, PRS transferability outside of European ancestry groups has been poorly studied. Moreover, there has been a dearth of suitable data to validate the utility of PRSs across a range of ancestry groups (particularly Africans, where glaucoma prevalence is more than twice that in other populations^1^).

Given the difficulties in early diagnosis, and the lack of cost-effective screening approaches, a more accurate genetic test would help clinicians apply timely evidence-based treatments to lower intraocular pressure (such as selective laser trabeculoplasty or topical medication^23,24^). It would also be highly beneficial for clinicians to understand which patients are at the highest risk of blindness, and may benefit more from earlier incisional surgical interventions which are highly efficacious^25^, but carry a risk of serious complications. In this work, we derive a glaucoma PRS with improved ability to stratify individuals and predict absolute risk across a wide range of ancestries. We also show this PRS is available for immediate clinical use, to predict more rapid disease progression, requirement for treatment in glaucoma suspects, and need for treatment escalation in established cases.

## METHODS

### PRS DEVELOPMENT

Glaucoma case-control GWAS summary statistics on an unprecedented scale were provided by 23andMe, Inc.; 4.5 million individuals, including 135,000 cases, from European (EUR), African (AFR) and Hispanic/Admixed American (AMR) groups. This was supplemented by existing GWAS summary statistics on glaucoma and its related traits (**Figure 1**, see also **Appendix**). The MTAG (version 1.0.8)^26^ multitrait approach accounts for any sample overlap between input datasets and leverages the genetic correlation between glaucoma and its related traits to maximize power; the input traits for the multitrait GWAS comprised a total of 297,871 glaucoma cases/6,524,164 controls, 159,452 individuals with intraocular pressure measures, 111,724 individuals with vertical cup-to-disc ratio measures and 54,968 cases of ocular hypertension/486,639 controls without ocular hypertension. A key feature of MTAG is that it outputs estimates for every marker in terms of each input trait and here we selected glaucoma risk (odds ratio) as the output trait. The GWAS output was used to construct a PRS for each ancestry group using SBayesRC (version v0.2.3) ^27^ (alternative methods did not improve performance, **Appendix Results**), with the per-ancestry PRSs then used to construct a single multi-ancestry PRS. The weights for combining each per-ancestry PRS into a single multi-ancestry PRS were estimated using a non-overlapping case-control dataset (75% of the NEI European case-control dataset, **Figure 1**). The weighted PRS (7 million SNPs with non-zero weights) was then adjusted for principal components of ancestry before being validated in non-overlapping datasets from the major ancestry groups.

**Figure 1:**
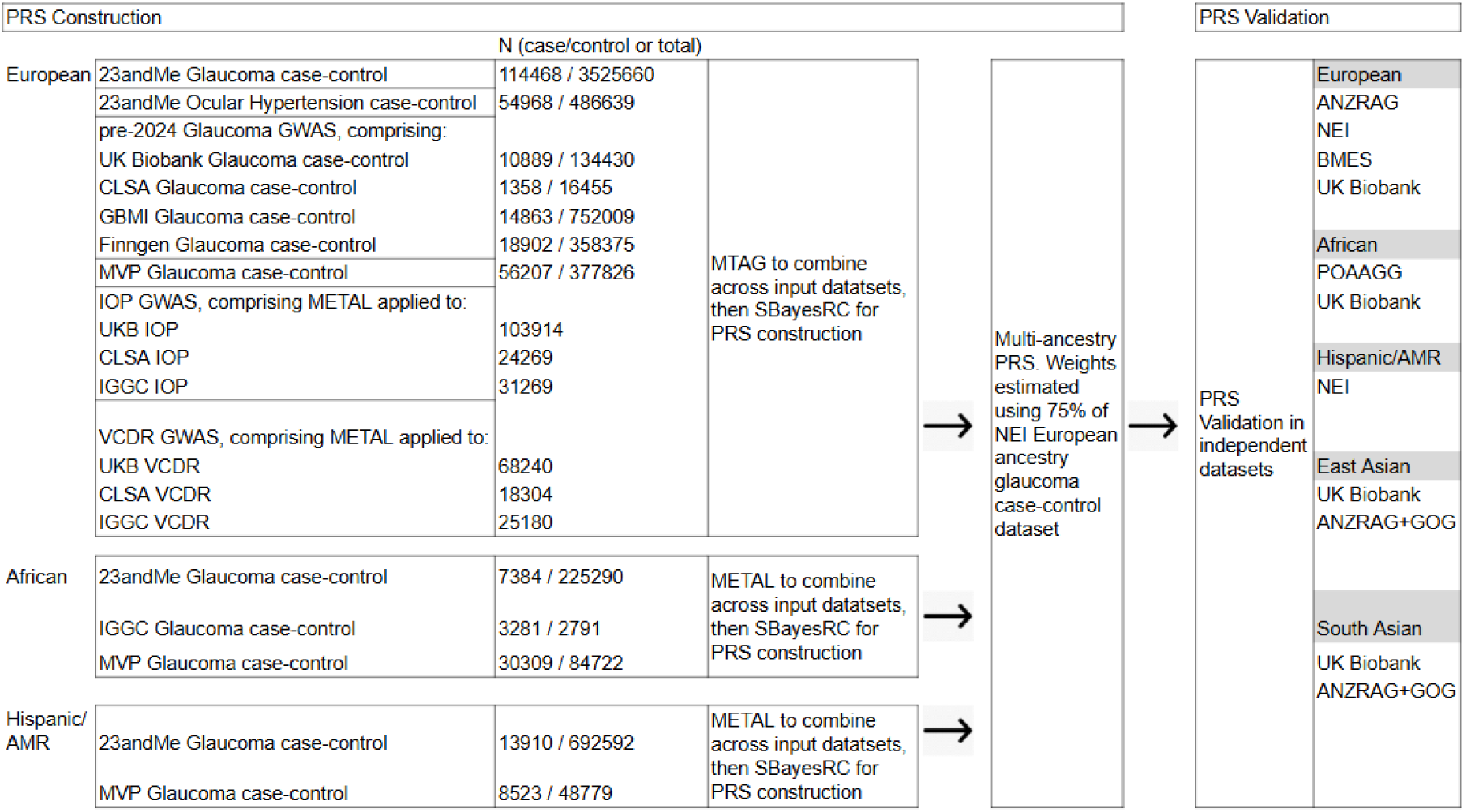
Composition and ancestry background of the cohorts used to generate and validate the glaucoma polygenic risk score. Ancestries are European (EUR), African (AFR), East Asian (EAS), South Asian (SAS), Admixed American/Hispanic (AMR). UKB=UK Biobank, BMES=Blue Mountains Eye Study, ANZRAG=Australian & New Zealand Registry of Advanced Glaucoma, GOG=Genetics of Glaucoma study, NEI=National Eye Institute Glaucoma Human Genetics Collaboration Heritable Overall Operational Database, POAAGG=Primary Open-Angle African American Glaucoma Genetics, CLSA=Canadian Longitudinal Study on Aging, GBMI=Global Biobank Meta-analysis Initiative, MVP=Million Veteran Program. UKB and CLSA glaucoma case-control meta-analysed using METAL, then combined with GBMI and Finngen using MTAG. The traits intraocular pressure (IOP) and vertical cup-to-disc ratio (VCDR) are highly correlated with glaucoma and improve power. All validation datasets are non-overlapping with data used in PRS construction and PRS weighting. For the NEI European ancestry dataset, the 25% of the dataset not used to estimate PRS weights was used as a validation set. Validation set sample sizes are in **Table 1**.

**Table 1.**
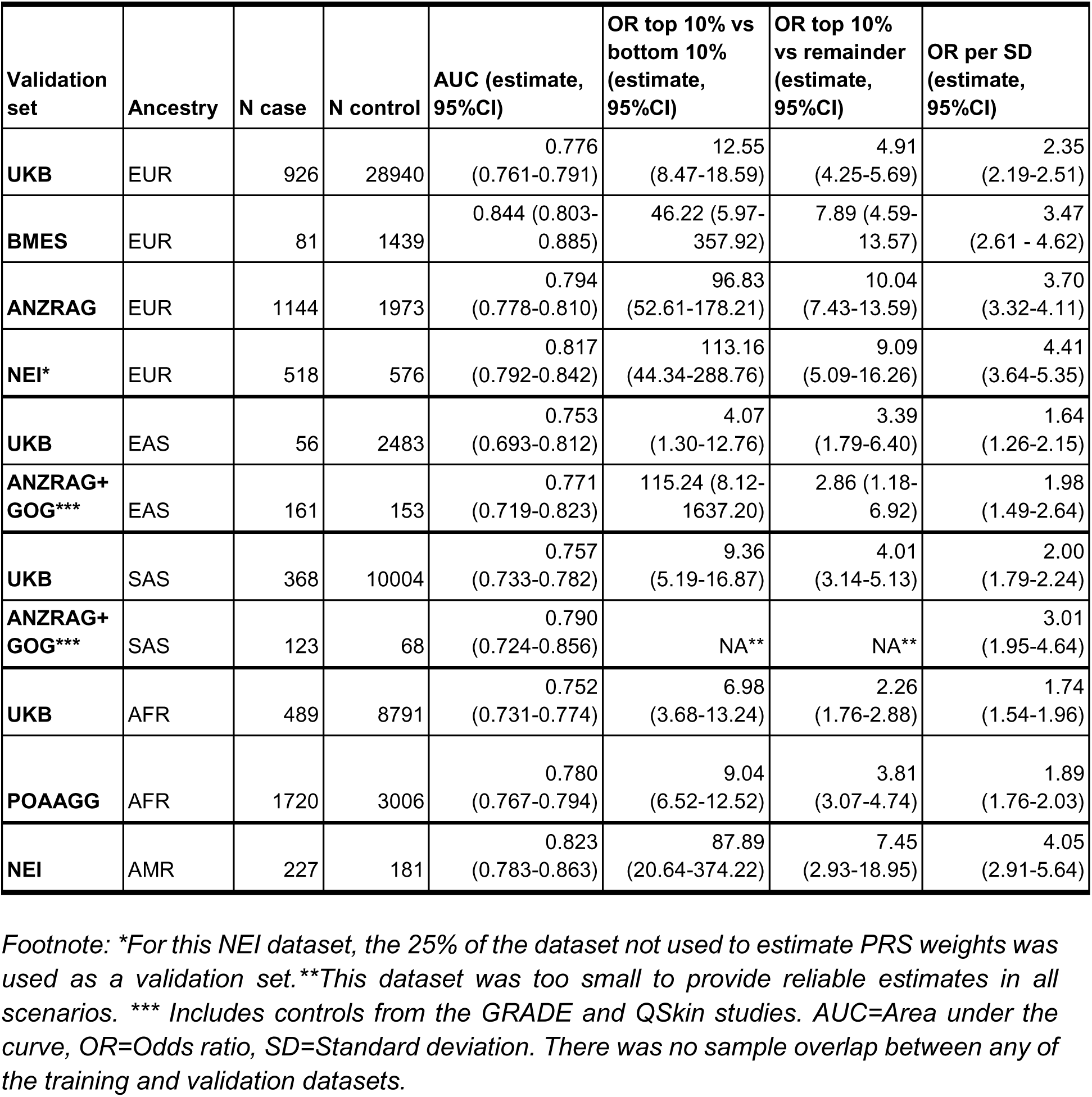
Predictive performance of the glaucoma polygenic risk score across independent cohorts with different ancestry backgrounds.

| Validation set | Ancestry | N case | N control | AUC (estimate, 95%CI) | OR top 10% vs bottom 10% (estimate, 95%CI) | OR top 10% vs remainder (estimate, 95%CI) | OR per SD (estimate, 95%CI) |
| --- | --- | --- | --- | --- | --- | --- | --- |
| **UKB** | EUR | 926 | 28940 | 0.776<br>(0.761-0.791) | 12.55<br>(8.47-18.59) | 4.91<br>(4.25-5.69) | 2.35<br>(2.19-2.51) |
| **BMES** | EUR | 81 | 1439 | 0.844 (0.803-0.885) | 46.22 (5.97-357.92) | 7.89 (4.59-13.57) | 3.47<br>(2.61 - 4.62) |
| **ANZRAG** | EUR | 1144 | 1973 | 0.794<br>(0.778-0.810) | 96.83<br>(52.61-178.21) | 10.04<br>(7.43-13.59) | 3.70<br>(3.32-4.11) |
| **NEI\*** | EUR | 518 | 576 | 0.817<br>(0.792-0.842) | 113.16<br>(44.34-288.76) | 9.09<br>(5.09-16.26) | 4.41<br>(3.64-5.35) |
| **UKB** | EAS | 56 | 2483 | 0.753<br>(0.693-0.812) | 4.07<br>(1.30-12.76) | 3.39<br>(1.79-6.40) | 1.64<br>(1.26-2.15) |
| **ANZRAG+**<br>**GOG\*\*\*** | EAS | 161 | 153 | 0.771<br>(0.719-0.823) | 115.24 (8.12-1637.20) | 2.86 (1.18-6.92) | 1.98<br>(1.49-2.64) |
| **UKB** | SAS | 368 | 10004 | 0.757<br>(0.733-0.782) | 9.36<br>(5.19-16.87) | 4.01<br>(3.14-5.13) | 2.00<br>(1.79-2.24) |
| **ANZRAG+**<br>**GOG\*\*\*** | SAS | 123 | 68 | 0.790<br>(0.724-0.856) | NA** | NA** | 3.01<br>(1.95-4.64) |
| **UKB** | AFR | 489 | 8791 | 0.752<br>(0.731-0.774) | 6.98<br>(3.68-13.24) | 2.26<br>(1.76-2.88) | 1.74<br>(1.54-1.96) |
| **POAAGG** | AFR | 1720 | 3006 | 0.780<br>(0.767-0.794) | 9.04<br>(6.52-12.52) | 3.81<br>(3.07-4.74) | 1.89<br>(1.76-2.03) |
| **NEI** | AMR | 227 | 181 | 0.823<br>(0.783-0.863) | 87.89<br>(20.64-374.22) | 7.45<br>(2.93-18.95) | 4.05<br>(2.91-5.64) |

Footnote: \*For this NEI dataset, the 25% of the dataset not used to estimate PRS weights was used as a validation set. \*\*This dataset was too small to provide reliable estimates in all scenarios. \*\*\* Includes controls from the GRADE and QSkin studies. AUC=Area under the curve, OR=Odds ratio, SD=Standard deviation. There was no sample overlap between any of the training and validation datasets.

### VALIDATION SETS - DISEASE RISK / CLINICAL PARAMETERS / FAMILY HISTORY

Multi-ancestry validation sets for disease risk were drawn from several studies (**Table 1**, **Appendix**).

We examined a series of clinical parameters in 1) PROGRESSA, a prospective longitudinal study with genetic data on glaucoma suspects and early-stage glaucoma cases, 2) ANZRAG a registry of established glaucoma cases and 3) two UK glaucoma cohorts (**Figure 3, 4, Appendix**).

We also assessed the association between the PRS and family history in the Genetics of Glaucoma and ANZRAG studies (**Appendix**).

### STATISTICAL ANALYSIS

#### PRS CONSTRUCTION

The input GWAS summary statistics (**Figure 1**) were meta-analysed using METAL (2011-03-25 release)^28^ when the input dataset was for the same trait (case-control or IOP or VCDR) and there was no sample overlap. Where there was sample overlap (for UKB/GBMI/Finngen case-control data) or where the traits being combined were different (case-control combined with quantitative traits), datasets were meta-analysed using MTAG. For the EUR group, the MTAG output for glaucoma risk was adjusted for corneal thickness (**Appendix**).

Following the GWAS meta-analysis, a PRS was constructed for each ancestry group using SBayesRC, with the per-ancestry PRSs then used to construct a weighted multi-ancestry PRS. The weights for the weighted PRSs were estimated using logistic regression (using R software, v3.4.1^29^) in a random subset (75%) of the NEI European ancestry case-control dataset. The independent validation set for the NEI Europeans was the remaining 25% of the NEI dataset (all other PRS validation sets were entirely separate studies, **Figure 1**). The 3 input PRSs were then linearly combined into a single multi-ancestry PRS.

The multi-ancestry PRS was then normalized for the first 5 principal components (PCs) of ancestry. We computed PCs on the 1000 Genomes Phase 3 dataset (comprising 2500 EUR, AFR, EAS, SAS, AMR individuals) and projected each individual requiring an ancestry adjusted PRS into the genetic ancestry space. We performed both mean and variance normalization, resulting in a PRS value for each individual which is standardized to mean 0, variance 1 within their genetic ancestry group. In principle, after normalization, the PRS can be used to risk stratify everyone relative to their ancestry background, including people who don’t fall close to one of the five 1000G super-populations based on genetic PCs. In practice, for our validations here, we focus on estimating the OR per SD within validation sets which are genetically similar to either EUR, AFR, EAS, SAS or AMR.

#### ASSOCIATION BETWEEN PRS AND PHENOTYPES

PRS estimates for each person in the validation samples were computed using the multi-ancestry PRS weights in PLINK v1.90b7^30^. Association between the PRS and phenotypes was conducted using logistic (disease risk, clinical outcomes other than incident surgery and remaining retinal nerve fibre layer in PROGRESSA), cox proportional hazard (incident surgery in PROGRESSA), linear regression (number of affected relatives), linear mixed-effects regression adjusting for inter-eye correlation (remaining retinal nerve fibre layer in PROGRESSA) models in R. “pROC” (version 1.18.5) was used to calculate the area under the curve (AUC) for binary traits^31^. Statistical analysis and plots were generated using R version 4.5.0.

To predict absolute risk, the probability of being a case was estimated from a logistic regression with age, sex and PRS fitted to validation sets which represented population samples (UK Biobank, BMES). We used this same logistic regression applied to BMES to determine the relationship between PRS percentile and disease age at onset (association between PRS and the average years later the glaucoma prevalence reaches 3%) and predicted risk over the next decade.

There was no sample overlap between any of the training and validation datasets. All reported p-values are from two-sided tests.

## RESULTS

### PRS PERFORMANCE - GLAUCOMA RISK

Our novel PRS showed marked improvements in OAG risk prediction performance in held out validation sets (**Table 1**). In the Australian ANZRAG European ancestry subset, individuals in the top 10% PRS had 10 fold increased risk (OR=10.0) relative to the remainder (OR was 4.2 for our previously published PRS^13^; the AUC also increased from 0.696 to 0.794, **Table S1**). Similar performance was seen in a subset of the US NEIGHBOR data not used for PRS development (OR=9.1 for top 10% versus remainder in European ancestry individuals; OR 7.5 in Hispanic/AMR). The PRS also offered exceptional differentiation between high (top 10%) and low (bottom 10%) risk with an OR of ∼100 (OR was 15 for our previously published PRS^13^) in three datasets (ANZRAG EUR, NEI EUR, NEI AMR).

In other ancestry groups, the novel PRS had similar performance to that seen in Europeans using our previous PRS^13^ (OR =4.0, OR=3.4 and OR=3.8 for top 10% versus the remainder in South Asians in UK Biobank, East Asians in UK Biobank and African Americans in the POAAGG study, respectively). Reconstructing the PRS without the large 23andMe input datasets reduced the performance of the PRS (OR=6.6 and 3.0 for top 10% relative to the remainder for ANZRAG EUR and POAAGG AFR datasets, respectively, **Table S1).** We constructed our PRS using the 3 largest populations with 23andMe GWAS results available (African American, Latino and European). We tested 2 alternative approaches. First we added East Asians but this did not meaningfully change PRS performance. Second we re-weighted the per-ancestry PRSs using an African instead of a European dataset. Neither approach meaningfully improved performance (**Table S2**, **Appendix Results**).

Some validation studies had IOP available on most cases and controls and we used this to assess prediction improvement when adding the PRS to a model with age, sex, PCs and IOP; in the NEIGHBOR study the AUC increased from 0.63 to 0.82 in EUR and from 0.68 to 0.83 in Hispanic/AMR. In the POAAGG study the AUC increased from 0.74 to 0.79 in AFR. For all of these datasets, after accounting for IOP, individuals with a top 10% PRS had strongly increased risk relative to the remainder of the population (EUR OR 7.93, AMR OR 7.11, AFR OR 4.10, **Table S3**).

The novel PRS effectively stratified an unselected population-based and fully examined Australian cohort (Blue Mountains Eye Study, BMES) for age at onset, with those in the top 10% PRS reaching 3% glaucoma prevalence on average >25 years earlier than those in the bottom 10% (**Figure 2, Panel 1**). **Figure 2, Panel 2** also shows the predicted absolute risk by PRS percentiles in BMES (based on models in **Table S4**). Those in the highest percentiles reach very high prevalence, whilst those at the low percentiles are very rarely affected; in the phase of BMES we use here (BMES3), where the age range is 60-97 (median 74), none of the 76 people in the bottom 5% PRS in BMES have glaucoma and only 1 is affected out of 152 people in the bottom 10% PRS (in contrast, in the top 10% PRS group, prevalence is 20%). The models in **Table S4** for BMES can also be used to inform the development of cost-effective PRS stratified screening guidelines; for example, a healthy European ancestry 65-year-old’s estimated glaucoma risk in the next 10 years is 10% for top 20% PRS individuals. In contrast, 10-year risk is only 0.4% or 1.0% for bottom 20% and 50% PRS individuals, respectively. The novel PRS also strongly separated people from a range of ancestry groups in UK Biobank into different absolute risk groups with increasing age and PRS percentiles (**Figure 2: Panels 3-6**). As expected, given the over-representation of Europeans in the training set, the separation between PRS percentiles is greatest in the independent European validation set. Among the four major groups in UK Biobank, African males with top 1% PRS reach the highest absolute risk (>35%) by age 80; this is due to the high baseline prevalence in Africans (∼2 times that of the other ancestries).

**Figure 2:**
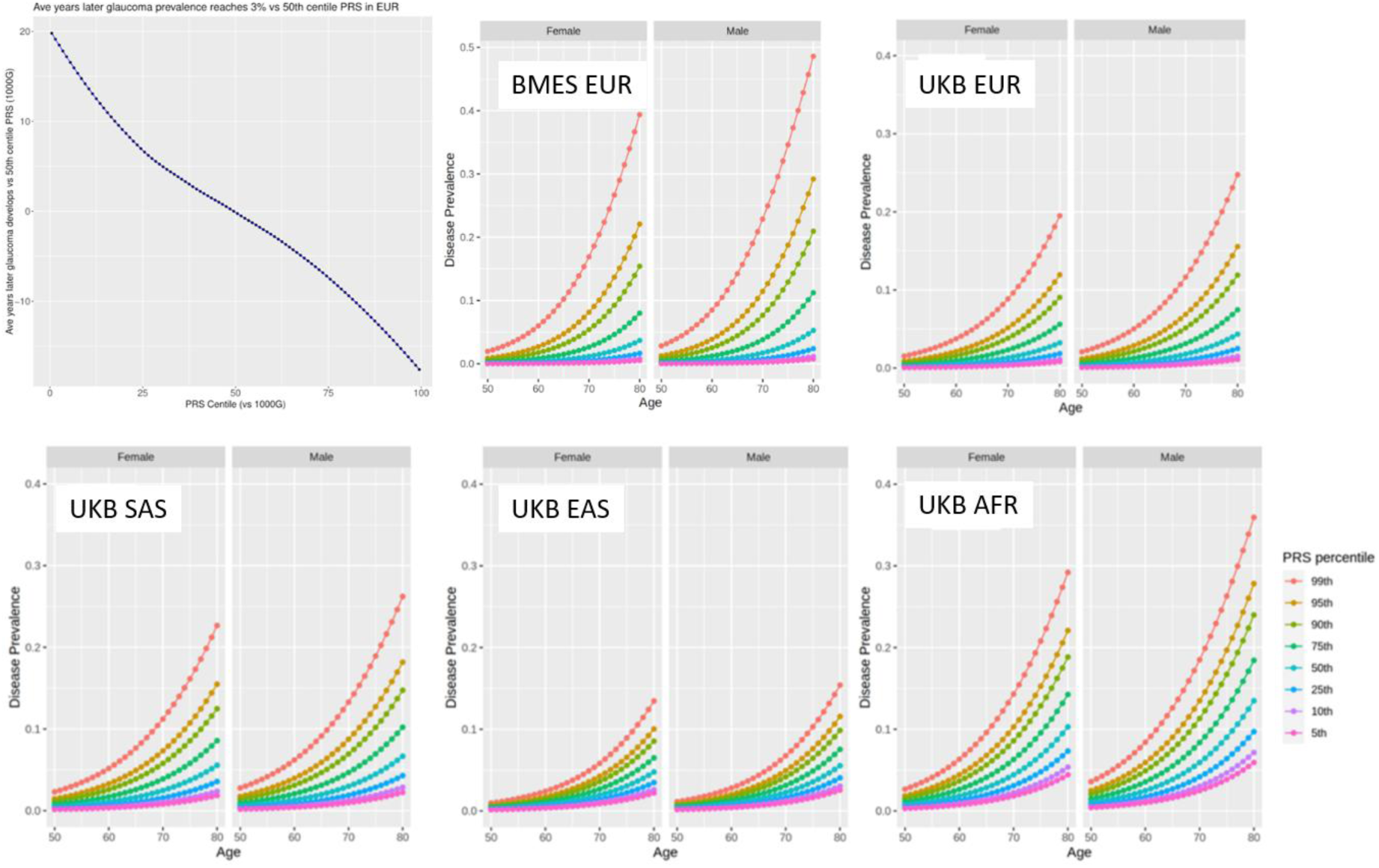
PRS and age. The first two panels show data from the population-based Blue Mountains Eye Study; the first shows the association between PRS and the average years later the glaucoma prevalence reaches 3%, relative to the 50th PRS percentile. The second panel shows the relationship between age and disease prevalence plotted by PRS percentile. Panels 3 to 6 shows the relationship between age and disease prevalence for each ancestry group in UK Biobank.

The new PRS and our previous PRS^13^ show moderate correlation (0.56 between percentile rank for each PRS in the GRADE cohort which comprises individuals from the general population, N=999 European ancestry individuals) although the ranking for any given person can change considerably. Importantly, when cases and controls are considered separately, it is clear that cases are being appropriately re-ranked; within the ANZRAG cohort, controls had a uniform distribution across PRS deciles for both the old and new PRS, but advanced glaucoma cases were heavily skewed toward high PRS deciles for the new PRS compared to the old PRS (**Figure S1**). Within ANZRAG cases, a much larger proportion of glaucoma patients are ranked highly for the new versus old PRS; half of cases are ranked within the top 12.7% and 25.0% PRS percentiles, for the new and old PRSs, respectively.

### CLINICAL IMPLICATIONS OF THIS PRS FOR OPEN-ANGLE GLAUCOMA

One of the hallmarks of early-stage glaucoma is retinal nerve fibre layer thinning. We quantified the longitudinal loss of retinal nerve fibre layer from baseline to latest follow-up for 2,448 eyes from 1,242 PROGRESSA participants with suspect or early manifest glaucoma. The novel PRS predicted the proportion of preserved retinal nerve fibre layer (**Figure 3A**), with an almost two-fold larger decrease per-decile compared with the previous PRS (P=0.044, **Figure S2**).

**Figure 3:**
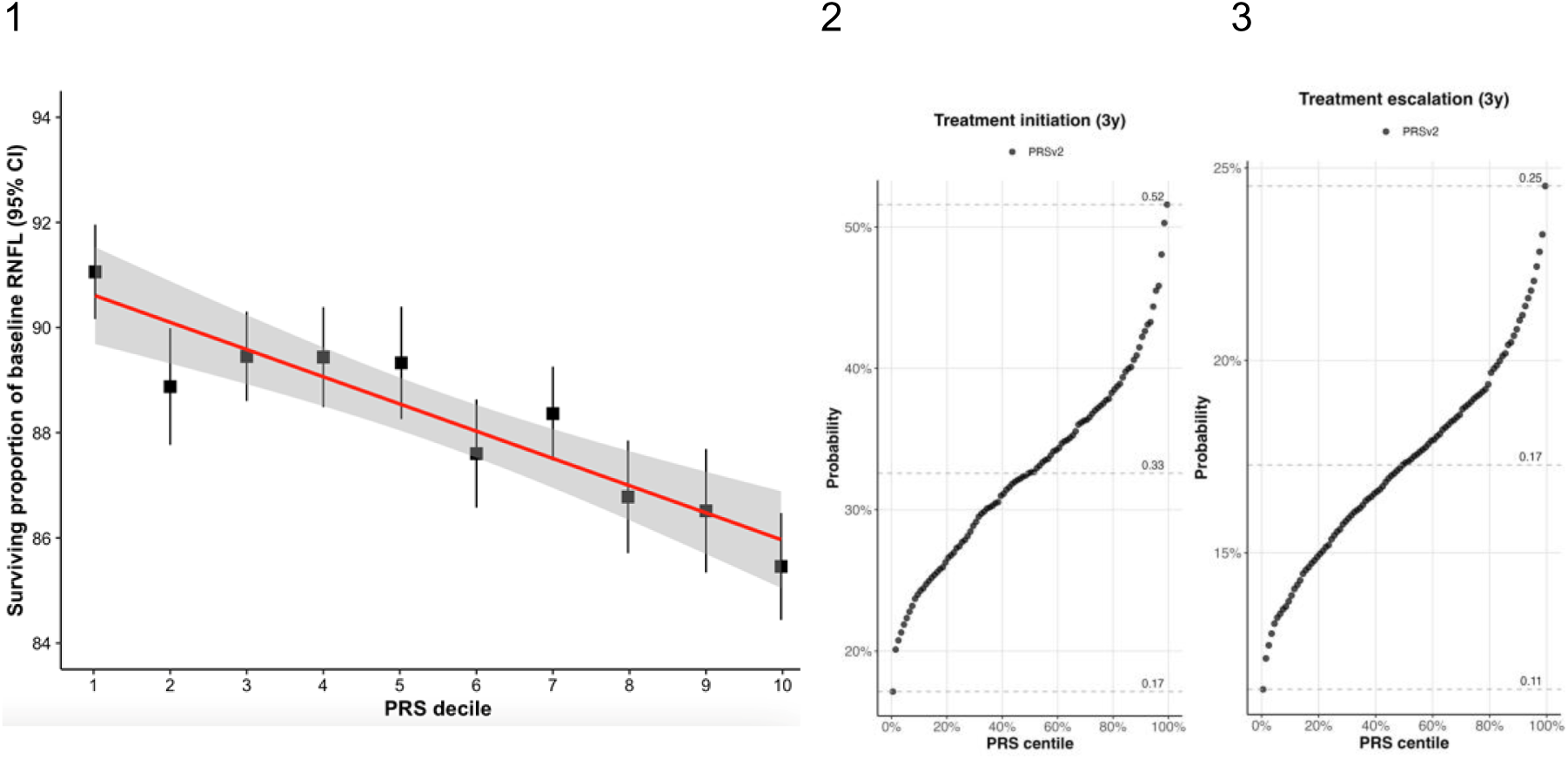
Impact of PRS on glaucoma progression and initiation and escalation of treatment. **Panel 1:** Proportion of preserved baseline retinal nerve fibre layer for N=2,448 eyes from N=1,242 PROGRESSA participants with suspect or early manifest primary open-angle glaucoma. The remaining retinal nerve fibre layer proportion is calculated for the most affected quadrant, as determined on optical coherence tomography scans at baseline and latest follow-up scan (mean duration of follow-up 7.91 years +/- 3.87). **Panels 2, 3:** Prediction of treatment initiation (N=473) or escalation (N=815) at 3 years. Treatment escalation events were defined as individuals who were on IOP-lowering treatment at enrollment, and received an additional topical medication, or SLT or surgical procedure.

Determining when to initiate treatment is difficult in glaucoma suspects. Though in such cases, the novel PRS was positively associated with treatment initiation within 3 years (OR per SD=1.39, 95%CI 1.14-1.70, P=0.0014) or treatment escalation within 3 years (OR per SD=1.21, 95%CI 1.00-1.45, P=0.047) (**Figure 3**). The novel PRS consistently performed better than the previous PRS, with particularly large improvement for treatment escalation (**Figure S3**).

Selecting the optimal time to surgically intervene for people with progressing glaucoma can be difficult, and substantial clinical resources are devoted to ensuring patients would appropriately benefit, both in early-stage and in advanced disease. We first assessed if the PRS could predict incident surgery (glaucoma filtration surgery or glaucoma drainage device) in prospectively monitored PROGRESSA participants with suspected or early manifest glaucoma at baseline, when their treating clinician did not know the patient’s PRS profile (N=1215, **Figure 4A**). Top PRS decile individuals were significantly more likely to proceed to surgery than those in mid or low deciles (P<0.0001), with 24% of participants in the top decile requiring surgery by age 80, compared with 0% in the bottom decile. The old PRS (**Figure S4**) showed less separation than the new PRS. Patients in the bottom PRS decile (where no individuals required surgery) are good candidates for reduced intensity of surveillance, while those in the top decile may benefit from earlier referral to ophthalmology specialists. The PRS was also associated with need for surgery in advanced glaucoma in cohorts from Australia (P=0.0004) and Liverpool UK (P=0.038) (**Figure 4**), with a similar, albeit not significant, trend in a Southampton UK cohort (**Figure S5**, **Table S5**).

**Figure 4:**
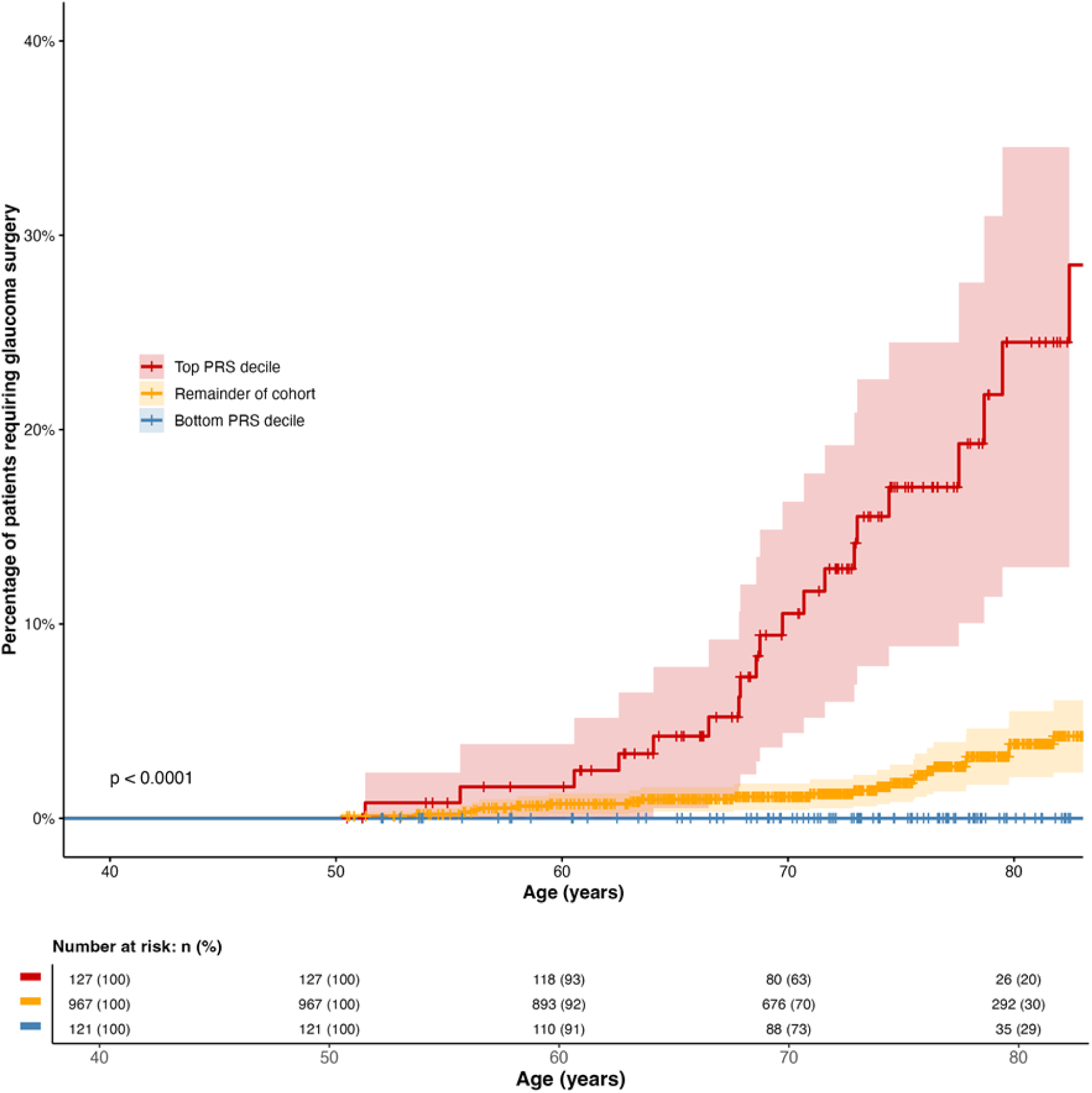
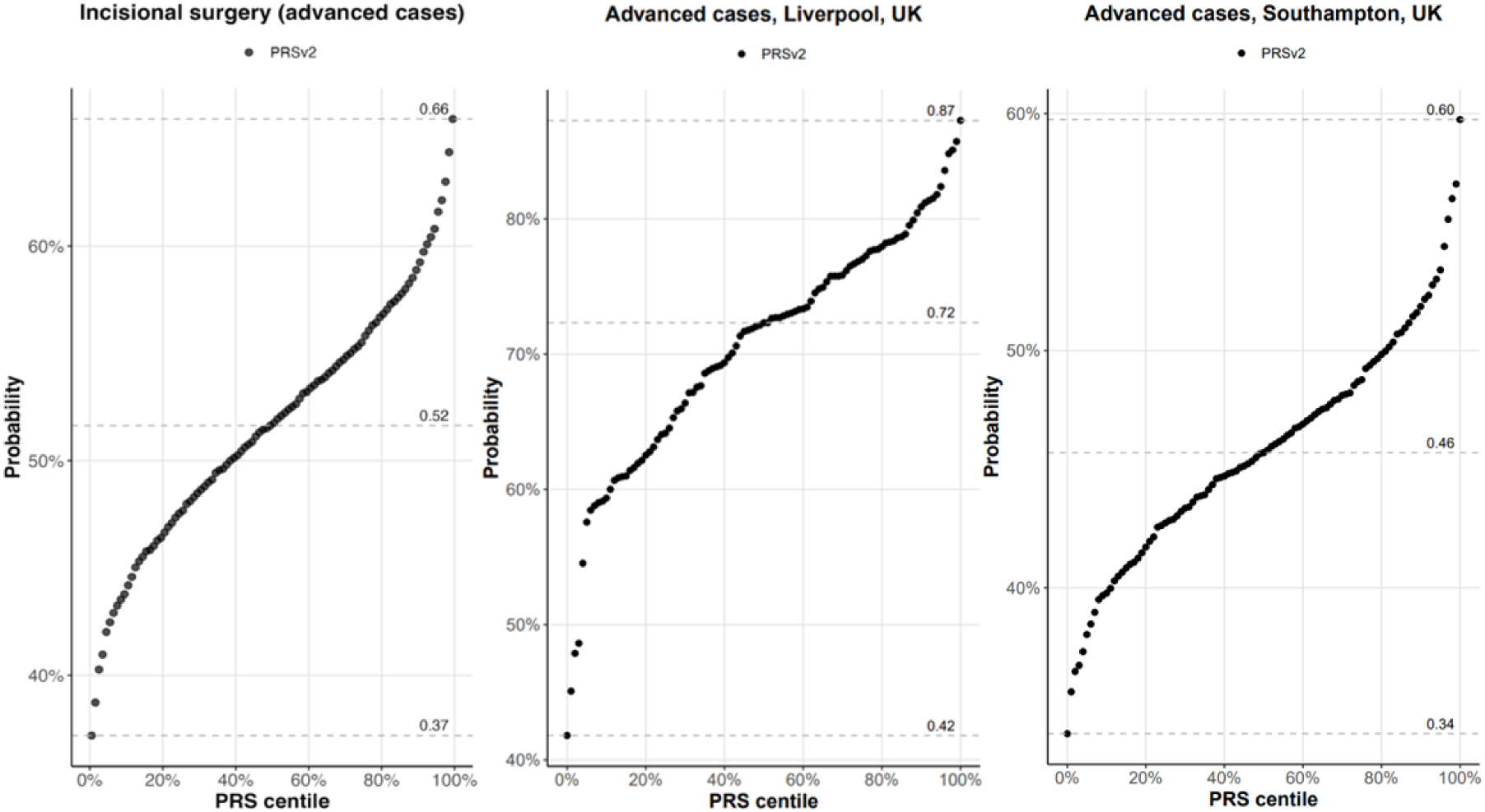
PRS predicts need for surgery in different clinical contexts. Panel 1: Prediction of incident surgery in a prospective study of suspected or early manifest glaucoma (PROGRESSA, N=1215, Cox proportional hazards model, P<0.0001). Survival was defined at age of first incisional surgery, in the worst affected eye. Cases were censored at the age of most recent follow-up. PRS is expressed as a decile internally derived from the PROGRESSA cohort. Red represents participants with a top decile PRS, blue represents participants with a bottom decile PRS, with the remaining individuals shown in yellow. Solid lines represent the median survival of the group, with 95% CI shaded in the respective color. Results shown are for the new PRS; results for the old PRS are shown in **Figure S4**. Panels 2-4: Association of PRS with need for surgery (logistic regression) in advanced glaucoma in ANZRAG (N=1434, P=0.000438, Panel 2), Liverpool (N=95, P=0.038, Panel 3) and Southampton (N=299, P=0.168, Panel 4).

In glaucoma cases from two studies the PRS predicted the probability of having a first-degree relative and the number of first-degree relatives, with consistent improvements for the novel PRS. In the Genetics of Glaucoma Study, for any first-degree relatives vs not (n=1698), the OR per 1 SD increase in PRS was 1.26 (95% confidence interval 1.14-1.40), P=1.3e-05. In the ANZRAG cohort (N=1565), the analogous OR per SD was smaller (OR 1.12, 95% confidence interval 1.08-1.16, P=5.0e-06), perhaps because the ANZRAG clinic based patients were less aware of their family history than Genetics of Glaucoma Study participants who represent a community based cohort who opted in to the study. The number of first-degree relatives increased by 0.11 (standard error 0.023) per 1 SD increase in PRS, P=1.5e-06 in Genetics of Glaucoma, with similar results in the ANZRAG cohort (**Figure S6**). Taken together, these results reinforce the fact that in addition to persons with very high PRS values being more likely to require treatment, they are also more likely to have family members with glaucoma (many of whom will be undiagnosed).

## DISCUSSION

We developed a glaucoma PRS with improved multi-ancestry performance, and offer robust evidence for direct clinical utility. The PRS enables high-risk stratification to identify those patients more likely to require closer monitoring for treatment escalation and glaucoma surgery before further irreversible vision loss occurs. The PRS also improves low-risk stratification, providing a foundation for less intensive monitoring and resource rationalisation for genetically lower-risk glaucoma suspects who are otherwise clinically ambiguous. The improved performance for both highest and lowest risk groups across the major ancestries offers a readily-available tool to develop cost-effective population screening^32^.

The large discovery sample size enabled a clear step change in PRS performance, with sample size key to performance, outweighing concerns about less accurate phenotyping in some datasets. The biggest contributor to our increased sample size was the increased number of glaucoma cases (∼300,000 for this version versus ∼8,000 in our previous work^13^), although we also harnessed larger sample sizes for GWASs of intraocular pressure, vertical cup to disc ratio (with AI derived rather than clinician derived phenotypes^33^), as well as new data on ocular hypertension. In European ancestry samples, the predominant ancestry in the training set, persons with top 10% PRS are at 10-fold increased risk relative to the rest of the population in independent well characterised cohorts (compared with ∼4-fold increased risk for our previous PRS^13^). In all ancestries, the new PRS offers a greater degree of risk stratification than is currently possible for any other *common* (>1% prevalence) complex disease, where OR per SD values are typically 1.5-2.2 for Europeans (compared with e.g. 4.4 in the NEI EUR cohort here) and lower for other ancestries. Indeed, while performance is strongest in Europeans, our glaucoma PRS shows similar or better performance in non-Europeans (OR per SD 1.64 to 3.01, **Table 1**) than most PRSs do for <u>any</u> ancestry for most other common complex diseases. The improved performance of our new PRS across different ancestries addresses previous health inequities highlighted by the US National Society of Genetic Counsellors and the Human Genetics Society of Australasia^34,35^.

While the new PRS shows some correlation with the old PRS, it is much better at ranking true cases highly. When benchmarked against general population PRS percentiles, 50% of cases in the ANZRAG cohort ranked in the top 12.7% of the new PRS (in contrast, the analogous figure for the old PRS is 25%). While population-wide screening for glaucoma (based on conventional risk factors such as IOP) is not thought to be cost-effective, previous studies have stressed that identifying subsets of the population at high risk would be key to making focused screening effective. Given the marked improvement in PRS performance (over and above IOP, **Table S3**) — the high case OR in the top decile of NEI and the near-zero incident glaucoma (despite a fully examined cohort) in the BMES bottom decile — future studies should fully evaluate the health economics of PRS based screening.

The new PRS offers strong risk stratification as age increases in population wide cohorts, with a 25-year difference in average age of onset between high and low PRS groups in an Australian population-based cohort with high-quality ocular phenotypes. We also provide predicted absolute risk estimates by age and PRS percentile for the four major ancestry groups in UK Biobank. Although UK Biobank enables us to estimate absolute risk across multiple ancestry groups, as a community-based cohort, not all glaucoma cases in UK Biobank will have been detected (about half of all cases are undiagnosed^36^), which means our absolute risk estimates underestimate the true risk (especially for early stage disease which is often undiagnosed). A further limitation is that, unlike BMES ^36^, UK Biobank is not representative of the general population. For European ancestries, we were able to estimate absolute risk by PRS percentile in BMES where all participants were examined for glaucoma. The higher quality glaucoma phenotype in BMES led to larger effect sizes (OR per SD for European ancestry individuals in BMES and UKB being 3.5 and 2.4, respectively), with this leading to larger absolute risk values for high PRS percentiles in BMES. The UKB OR per SD was also larger (2.5) if screened healthy controls were used instead of the 28940 controls in Table 1 (which comprise all non-cases in the ∼10% of UKB we reserved to validate the PRS). In the future, it would be advantageous to have population-based cohorts with full eye examinations from a range of ancestries to enable better estimation of absolute risk in these groups. As well as improving stratification for disease risk, our new PRS also predicted clinical outcomes in glaucoma suspects and in manifest glaucoma cases, with improvements in accuracy relative to a previous PRS.

Overall, our multi-ancestry PRS predicts glaucoma risk well in all ancestry groups. We adjusted for ancestry using principal components (PC), whereby the PRS was rendered on a standardized scale in each of our validation cohorts (e.g. for each ancestry group we can directly map each individual to a specific risk percentile). For the ancestries represented in our validation cohorts in **Table 1**, the OR per SD for each ancestry enables computation of a specific relative risk for a person, given their PRS risk percentile. Using a single PC-adjusted PRS for all ancestries has the advantage of allowing our PRS to be directly applicable to admixed individuals, although for mixed ancestry groups further work would be beneficial to determine the optimal OR per SD scaling for such individuals.

This PRS efficiently identifies individuals at very high-risk of developing glaucoma as well as an accelerated escalation of IOP-lowering treatments including surgical intervention. Risk profiling with the enhanced PRS will enable earlier screening and timely treatment of high-risk individuals, with reduced screening and monitoring costs in those at low risk. This PRS offers more discrimination, in terms of OR per SD, than any major *common* complex disease. This unprecedented ability to predict risk for glaucoma provides a foundation for more intensive monitoring and treatment escalation of high risk patients, and resource rationalisation for lower risk patients. It also provides a currently available, rapid turnaround, accredited instrument to develop cost-effective screening programs for the undiagnosed 2.4 million Americans^37^ at risk of irreversible vision loss.

## Acknowledgments

This work was conducted using the UK Biobank Resource (application number 25331) and publicly available data from the International Glaucoma Genetics Consortium. The UK Biobank was established by the Wellcome Trust, Medical Research Council (UK), Department of Health (UK), Scottish Government, and Northwest Regional Development Agency. It also had funding from the Welsh Assembly Government, British Heart Foundation, and Diabetes UK. The eye and vision dataset has been developed with additional funding from The NIHR Biomedical Research Centre at Moorfields Eye Hospital and the UCL Institute of Ophthalmology, Fight for Sight charity (UK), Moorfields Eye Charity (UK), The Macula Society (UK), The International Glaucoma Association (UK) and Alcon Research Institute (USA). This work uses data provided by patients and collected by the NHS as part of their care and support.

This work was supported by grants from the National Health and Medical Research Council (NHMRC) of Australia (#1107098; 1116360, 1116495, 1023911, 1150144), the Ophthalmic Research Institute of Australia, and the BrightFocus Foundation. SM, JEC, DAM, PG and AWH are supported by NHMRC grants (Fellowship/Investigator/Program), OMS by the Snow Medical Research Foundation (Grant No. PF2019-040), DAM by Glaucoma Australia, and Telethon. We thank David Whiteman and Catherine Olsen for coordinating all aspects of the QSKIN study.

APK is supported by a UK Research and Innovation Future Leaders Fellowship (MR/Y033930/1), an Alcon Research Institute Young Investigator Award and a Lister Institute for Preventive Medicine Award. This research was supported by the NIHR Biomedical Research Centre at Moorfields Eye Hospital and the UCL Institute of Ophthalmology.

We would like to thank the research participants and employees of 23andMe for making this work possible.

The following members of the 23andMe Research Team contributed to this study:

Stella Aslibekyan, Adam Auton, Elizabeth Babalola, Robert K. Bell, Jessica Bielenberg, Ninad S. Chaudhary, Zayn Cochinwala, Sayantan Das, Emily DelloRusso, Payam Dibaeinia, Sarah L. Elson, Nicholas Eriksson, Chris Eijsbouts, Teresa Filshtein, Pierre Fontanillas, Davide Foletti, Will Freyman, Zach Fuller, Julie M. Granka, Chris German, Éadaoin Harney, Alejandro Hernandez, Barry Hicks, David A. Hinds, M. Reza Jabalameli, Ethan M. Jewett, Yunxuan Jiang, Sotiris Karagounis, Lucy Kaufmann, Matt Kmiecik, Katelyn Kukar, Alan Kwong, Keng-Han Lin, Yanyu Liang, Bianca A. Llamas, Aly Khan, Steven J. Micheletti, Matthew H. McIntyre, Meghan E. Moreno, Priyanka Nandakumar, Dominique T. Nguyen, Jared O’Connell, Steve Pitts, G. David Poznik, Alexandra Reynoso, Shubham Saini, Morgan Schumacher, Leah Selcer, Anjali J. Shastri, Jingchunzi Shi, Suyash Shringarpure, Keaton Stagaman, Teague Sterling, Qiaojuan Jane Su, Joyce Y. Tung, Susana A. Tat, Vinh Tran, Xin Wang, Wei Wang, Catherine H. Weldon, Amy L. Williams, Peter Wilton.

dbGaP data from accessions phs001312.v1.p1 and phs000238.v1.p1 were used.

The Primary Open-Angle African American Glaucoma Genetics (POAAGG) Study is supported by funding the National Eye Institute, Bethesda, Maryland (grant # 5R01EY023557-03) and the Department of Ophthalmology at the Perelman School of Medicine, University of Pennsylvania, Philadelphia, PA. Funds also come from the F.M. Kirby Foundation, Research to Prevent Blindness, The Paul and Evanina Bell Mackall Foundation Trust, and the National Eye Institute, National Institutes of Health, Department of Health and Human Services, under eyeGENETM contract Nos. HHSN260220700001C and HHSN263201200001C. The sponsor or funding organization had no role in the design or conduct of this research. The origin of the dataset is described in the following publication: Charlson ES, Sankar PS, Miller-Ellis E, Regina M, Fertig R, Salinas J, Pistilli M, Salowe RJ, Rhodes AL, Merritt WT, Chua M, Trachtman BT, Gudiseva HV, Collins DW, Chavali VR, Nichols C, Henderer J, Ying GS, Varma R, Jorgenson E, O’Brien JM. (2015) The Primary Open-Angle African-American Glaucoma Genetics (POAAGG) Study: Baseline Demographics. Ophthalmology 122:711-20.

The dataset(s) used for the analyses described in this manuscript were obtained from the NEI Glaucoma Human Genetics Collaboration (NEIGHBOR) Dataset under the dbGaP accession number phs000238. Funding support for NEIGHBOR was provided by the National Eye Institute (EY022305). We would like to thank NEIGHBOR participants and the NEIGHBOR Research Group for their valuable contribution to this research.

NEI R01EY022305 (JLW) supports the NEIGHBOR study, which evolved into the NEIGHBORHOOD Consortium. LRP and JLW are supported by NEI R01EY032559, which focuses on the clinical utility of a glaucoma polygenic risk score for glaucoma in regional biobanks. LRP receives support from The Glaucoma Foundation (NYC) to explore the clinical utility of the glaucoma polygenic risk score.

## Ethics Statement

This study was approved by the QIMR Berghofer Human Research Ethics Committee. In addition, relevant details for each of the participating cohorts are provided below: UK Biobank: The UK Biobank study was approved by the National Health Service National Research Ethics Service (ref. 11/NW/0382) and all participants provided written, informed consent to participate in the UK Biobank study. Information about ethics oversight in the UK Biobank can be found at https://www.ukbiobank.ac.uk/ethics/. 23andMe Inc: Participants provided informed consent and participated in the research online, under a protocol approved by the external AAHRPP-accredited IRB, Ethical & Independent Review Services (E&I Review).

## Resource Availability

UK Biobank data are available through the UK Biobank Access Management System at https://www.ukbiobank.ac.uk/. The dbGaP datasets phs001312.v1.p1 and phs000238.v1.p1 are available through the authorised access portal at https://dbgap.ncbi.nlm.nih.gov/. To facilitate PRS benchmarking, we will provide per-individual PRS values for the main validation cohorts including ∼20,000 glaucoma cases and controls across all ancestry groups (EUR, AMR, AFR, EAS, SAS). These comprise UKB samples (EAS, SAS) and dbGaP samples [NEIGHBOR (EUR, AMR) and POAAGG (AFR)]. We will provide on request the dbGaP per-individual PRS values for NEIGHBOR and POAAGG. Per-individual PRS values will be returned to UKB for distribution to other approved UKB researchers. The PRS is available for clinical use via Seonix Bio; to support academic engagement the PRS is available for research use via the Seonix Bio collaborative platform.

## Author’s Contributions

S.M. and J.E.C. designed the study. S.M., G.N., M.S., A.K., N.Q.L., J.S., P.G., V.T., P.N., S.S., J.M.G., O.M.S. analyzed the data. S.M., P.G., A.W.H., J.E.C., J.L.W., J.O., R.N.W., P.M., N.H., O.M.S. and D.A.M. obtained funding. S.M., A.K., J.S., P.G., V.T., E.S., P.N., S.S., J.M.G., T.A., J.L., S.L.G., P.R.H., A.P.K., L.R.P., J.L.W, J.O., R.N.W., C.E.W., A.L., P.M., D.A.M., A.W.H, O.M.S., J.E.C. contributed to data collection and contributed to genotyping. S.M. wrote the first draft of the paper. All authors contributed to the final version of the paper.

## Supplementary Appendix

### Supplementary Appendix Methods

#### Abbreviations

Admixed American/Hispanic (AMR)

African (AFR)

Area under the curve (AUC)

Australian & New Zealand Registry of Advanced Glaucoma (ANZRAG)

Blue Mountains Eye Study (BMES)

Canadian Longitudinal Study on Aging (CLSA)

East Asian (EAS)

European (EUR)

Genetics of Glaucoma study (GOG)

Genome-wide association study (GWAS)

Global Biobank Meta-analysis Initiative (GBMI)

Genetic Risk Assessment of Degenerative Eye Disease study (GRADE)

International Glaucoma Genetics Consortium (IGGC)

Intraocular pressure (IOP)

Meta-analysis helper (METAL - software package) Million Veteran Program (MVP)

Multiple trait analysis of GWAS (MTAG - software package)

National Eye Institute Glaucoma Human Genetics Collaboration Heritable Overall Operational Database (NEI)

Open-angle glaucoma (OAG)

Polygenic risk score (PRS)

Primary Open-Angle African American Glaucoma Genetics (POAAGG)

Progression Risk Of Glaucoma: RElevant SNPs with Significant Association study (PROGRESSA)

Retinal nerve fibre layer (RNFL)

Single-nucleotide polymorphisms (SNP) South Asian (SAS)

UK Biobank (UKB)

Vertical cup-disc ratio (VCDR)

#### INPUT DATASETS FOR PRS CONSTRUCTION

##### Training Set: 23andMe case-control

A glaucoma case-control GWAS was conducted separately in 3 groups: European (EUR), African (AFR), Admixed American/Hispanic (AMR) (Fig. 1). Cases were defined as those reporting glaucoma, excluding those with closed-angle glaucoma or other types of glaucoma. Controls were those without glaucoma. Additionally, a separate case-control genome-wide association study (GWAS) of ocular hypertension was run in the European group; cases report elevated intraocular pressure (IOP), controls did not have elevated IOP or glaucoma. Participants provided informed consent and volunteered to participate in the research online, under a protocol approved by the external AAHRPP-accredited Salus IRB (https://www.versiticlinicaltrials.org/salusirb).

The GWAS for glaucoma and for ocular hypertension was run using logistic regression with an additive model and sex, age, 10 principal components (PCs) of ancestry and genotyping platform as covariates. Unrelated individuals (identity-by-descent (IBD) sharing < 700 cM) were included for the GWAS. The genotyping, ancestry determination, and imputation (based on combining the Haplotype Reference Consortium plus internal sequencing data) are described in detail elsewhere.^1^

Training Set: Million Veterans Program (MVP) glaucoma case-control Previously published GWAS summary statistics for EUR, AFR and AMR genetically inferred ancestry groups were collated from MVP release 4^2^. Cases had open angle glaucoma; controls did not.

##### Training Set: Pre-2024 glaucoma case-control

These case-control datasets comprised previously published studies plus a version of UK Biobank curated for use in this study. We collated GWAS data from Finngen release 9 (glaucoma based on ICD codes H40-42, versus not). We used previously published primary open angle case-control data from the Global Biobank Meta-analysis Initiative^3^ and previously published data from Canadian Longitudinal Study of Aging (self reported glaucoma, versus not)^4^.

UK Biobank (UKB) is a population-based study of ∼500,000 people living in the United Kingdom.^5^ UK Biobank samples were genotyped with Affymetrix Axiom arrays and quality control procedures were reported previously.^5^ All participants provided informed written consent and the study was approved by the National Research Ethics Service Committee North West – Haydock. Here, we used only individuals with kinship relationships lower than 0.1. Cases were those who were either an ICD10 case, an ICD9 case, a self-reported glaucoma case (data field 6148), or a glaucoma case (data field 20002). Controls were those who were not a self-reported case, not an ICD10 or ICD9 case, didn’t have eye disease, and didn’t have any eye disease-related surgery. We identified 11,815 EUR cases and 139,742 EUR controls. Out of this set, we randomly held out a subset as a validation dataset to evaluate the prediction performance. To avoid sample overlap with persons with IOP measures, the held out set was selected to be ∼10% of persons without an IOP measurement.

##### Training Set: Intraocular pressure and certical cup-disc ratio (VCDR)

We used publicly available European ancestry GWAS summary statistics for IOP and for VCDR from European ancestry individuals from the International Glaucoma Genetic Consortium (IGGC)^6^ and from UK Biobank and the Canadian Longitudinal Study of Aging^7^.

#### VALIDATION SETS - DISEASE RISK

##### Validation set: UK Biobank

A subset of the UKB European ancestry samples were retained as an independent validation set. Additionally, UKB validation sets were formed from individuals of African, South Asian and East Asian ancestry (Sample sizes in Table 1). Cases were those who were either an ICD10 case, an ICD9 case, a self-reported glaucoma case (data field 6148), or a glaucoma case (data field 20002). To allow estimation of absolute risk, controls were those in the independent validation set who were not cases. For absolute risk estimation age in 2023 was defined as current age for controls, age at onset (data field 131186) for cases; for a small number of cases with missing age at onset, current age was used. Age, sex and 10 principal components were fitted as covariates.

##### Validation sets: Australia

For the European ancestry validation, primary open angle advanced glaucoma cases from the Australian & New Zealand Registry of Advanced Glaucoma (ANZRAG) were compared to controls^8^. Sex and 10 principal components were fitted as covariates (age was unavailable on controls). A further European ancestry validation set was drawn from the second follow-up of the Blue Mountains Eye Study (BMES3), an Australian population-based cohort study^9^; cases had diagnosed open angle glaucoma, while controls did not. Age, sex and 10 principal components were fitted as covariates.

For Australians of East Asian (EAS) and South Asian (SAS) ancestry, validation was performed with open angle glaucoma cases drawn from ANZRAG, with further (self report) glaucoma cases drawn from the Genetics of Glaucoma Study^10^; ancestry matched controls were drawn from the GRADE^11^ and QSkin studies^12^. Age, sex and 10 principal components were fitted as covariates. 999 participants of European ancestry from the Genetic Risk Assessment of Degenerative Eye Disease study (GRADE) study were also used to compute the correlation between the PRS percentiles for the current PRS and a previously published PRS^13^.

The ANZRAG European ancestry validation samples were genotyped on Illumina Omni1M or OmniExpress arrays^8^. GRADE samples were genotyped on Illumina GSA arrays and were also used to illustrate the difference in PRS rankings for the current PRS and a previously published PRS^13^. The Blue Mountains Eye Study was genotyped with Illumina Human610-Quad arrays^9^.

For the East Asian (EAS) and South Asian (SAS) validation sets, due to limited sample size we drew cases from both the Australian & New Zealand Registry of Advanced Glaucoma (ANZRAG) and Genetics of Glaucoma (GOG) studies. Ancestry matched controls were drawn from the QSkin and GRADE studies, with all samples genotyped on Illumina GSA arrays. The sample sizes for EAS were GOG: 72 cases, ANZRAG: 89 cases, QSkin: 131 controls, GRADE: 22 controls. The sample sizes for SAS were GOG: 57 cases, ANZRAG: 66 cases, QSKIN: 63 controls, GRADE: 5 controls.

##### Validation set: USA

For validation in Europeans and in Admixed Americans, POAG cases and healthy controls were drawn from the National Eye Institute Glaucoma Human Genetics Collaboration Heritable Overall Operational Database (NEIGHBORHOOD) dataset (phs000238.v1.p1). In Europeans the validation set was the 25% of the cohort not used in estimating PRS weights; in Admixed Americans all cases and controls were used. For validation in African ancestry individuals, POAG cases and healthy controls were drawn from the Primary Open-Angle African American Glaucoma Genetics (POAAGG) Study (phs001312.v1.p1). Age, sex and 10 principal components were fitted as covariates.

NEIGHBORHOOD samples were genotyped on Illumina Human660W arrays. POAAGG samples were genotyped on the Illumina Infinium Multi-Ethnic Genotyping Array (MEGA).

#### VALIDATION SETS - CLINICAL PARAMETERS

We examined a series of clinical parameters in Progression Risk Of Glaucoma: RElevant SNPs with Significant Association (PROGRESSA), a prospective longitudinal study with genetic data on glaucoma suspects and early-stage glaucoma cases. Participants were genotyped on Illumina HumanCoreExome and GSA arrays. We recorded the proportion of preserved baseline retinal nerve fibre layer (RNFL) for PROGRESSA participants; the remaining RNFL proportion was calculated for the most affected quadrant, as determined on optical coherence tomography scans at baseline and latest follow-up scan (N=2,448 eyes from N=1,242 participants). Data was also collated from clinical records for treatment initiation within 3 years (N=473) and treatment escalation within 3 years (N=815). Treatment escalation events were defined as individuals who were on IOP-lowering treatment at enrollment, and received an additional topical medication, or selective laser trabeculoplasty (SLT) or surgical procedure. Age and sex were fitted as covariates.

Furthermore, we also collated data on incident incisional surgery collated prospectively in N=1215 PROGRESSA participants with suspected or early manifest glaucoma at baseline; survival was defined at age of first incisional surgery, in the worst affected eye as determined by lowest visual field mean deviation (dB). Cases were censored at their age of most recent follow up; sex was fitted as a covariate. We also collated incisional surgery data from clinical records for advanced glaucoma cases from Australian & New Zealand Registry of Advanced Glaucoma (ANZRAG); N=1434 advanced cases, excluding known monogenic causes, with an age at diagnosis of ≥40 years. Age and sex were fitted as covariates. All ANZRAG and PROGRESSA participants included were of European ancestry.

We also collated surgery status (incisional or glaucoma filtration surgery) from POAG patients from two UK cohorts (Liverpool, N=95, and Southampton, N=299), as described previously^13^. Samples were genotyped on Illumina Infinium Global Screening Array-24 arrays.

Participants were of European ancestry and age, sex, 10 principal components and maximum recorded IOP were fitted as covariates.

#### VALIDATION SETS - FAMILY HISTORY

First degree relative status (yes/no) and number of affected first degree relatives was taken from clinical records for European ancestry advanced POAG cases from ANZRAG (N=1565 advanced POAG cases, excluding known monogenic causes, with an age at diagnosis of ≥40 years). Analogous data based on self report first degree relative status was taken from N=1698 European ancestry glaucoma cases from the Genetics of Glaucoma Study. Age, sex and 10 principal components were fitted as covariates.

For all validation datasets ancestry was genetically determined, with individuals categorized into EUR, AFR, EAS, SAS, AMR as defined in 1000 Genomes based on a K-means clustering method based on genetic principal components.

#### PRS CONSTRUCTION METHODS

Following meta-analysis of the input GWAS datasets (Figure 1), for the EUR group, the MTAG output for glaucoma risk (odds ratio, 23andMe glaucoma case-control), was adjusted for corneal thickness using a previously published corneal thickness GWAS^29^ via the mtCOJO method (as implemented in GCTA version v1.94.1; this effectively regresses out corneal thickness from the glaucoma case-control analysis) and then used for PRS construction. For the AFR and AMR groups, the METAL output was used for PRS construction (not corrected for corneal thickness as there are no very large corneal thickness GWASs in these ancestries).

#### EXPLORATORY PRS CONSTRUCTION METHODS

##### East Asian Training sets

We conducted an exploratory analysis to test a PRS made from East Asian datasets. We used MTAG (version 1.0.8) to combine 9715 glaucoma cases and 260245 controls from GBMI^3^, 6935 glaucoma cases and 39588 controls from IGGC^14^ and 1127 open angle glaucoma cases and 5402 controls from MVP^2^. A PRS was constructed by applying SBayesRC (version v0.2.3) to the East Asian MTAG output. The PRS was compared with our multi-ancestry PRS on the UKB EAS cohort. GWAS sample sizes in SAS populations were insufficient to build a PRS, although we did compare our multi-ancestry PRS with the EAS only PRS.

### Supplementary Results

We tested various PRS construction methods and found that SBayesRC offered the best risk prediction performance. In particular, the ability of SBayesRc to use 7 million SNPs in the PRS led to marked improvements in performance relative to approaches using only ∼1 million Hapmap3 SNPs. When we tested alternative approaches to create multi-ancestry PRSs which only use Hapmap3 (such as PRS-CSx), the performance for each ancestry group was worse than our approach using 7 million SNPs in a SBayesRC-derived PRS.

We constructed our PRS focusing on the 3 largest populations with 23andMe GWAS results available (African American, Latino and European). We tested (**Appendix**, **Table S2**) if a PRS made by applying SBayesRC to a GWAS meta-analysis of East Asian individuals from IGGC^14^, GBMI^3^ and MVP^2,3^ improved PRS performance in our East Asian validation set; the performance was worse, likely due to the much larger sample size in our three largest populations. We conducted exploratory analysis adding the smaller datasets of East Asian ancestry to the other three populations, although this did not improve the predictive performance in any of the tested datasets. For our multi-ancestry PRS, we estimated the weights using European ancestry individuals from the NEI study. We also tested estimating the weights in a subset of an African American ancestry datasets (POAAGG) but this did not substantially change performance when tested in the remainder of the POAAGG dataset.

Our main analysis adjusted the PRS for the first 5 PCs estimated from the 1000 Genomes superpopulations (AFR, AMR, EUR, EAS, SAS), with this adjusting for any broad population differences in the PRS distribution. To address a possible concern that adjusting for PCs may adversely affect the results, we also computed the OR per SD change without PC adjustment and the results were was essentially unchanged (e.g. in the ANZRAG+GOG EAS data the OR per SD was 1.99, compared with the 1.98 with PC adjustment, Table 1).

### Supplementary Appendix Tables

Excel spreadsheet contains all Tables (Table S1 - Table S4)

**Table S1 - Comparison between different PRSs**

**Table S2 - UKB and BMES PRS results**

**Table S3 - Effect of PRS on the need for surgery in advanced glaucoma**

**Table S4 - Asian PRS results**

### Supplementary Appendix Figures

**Figure S1:**
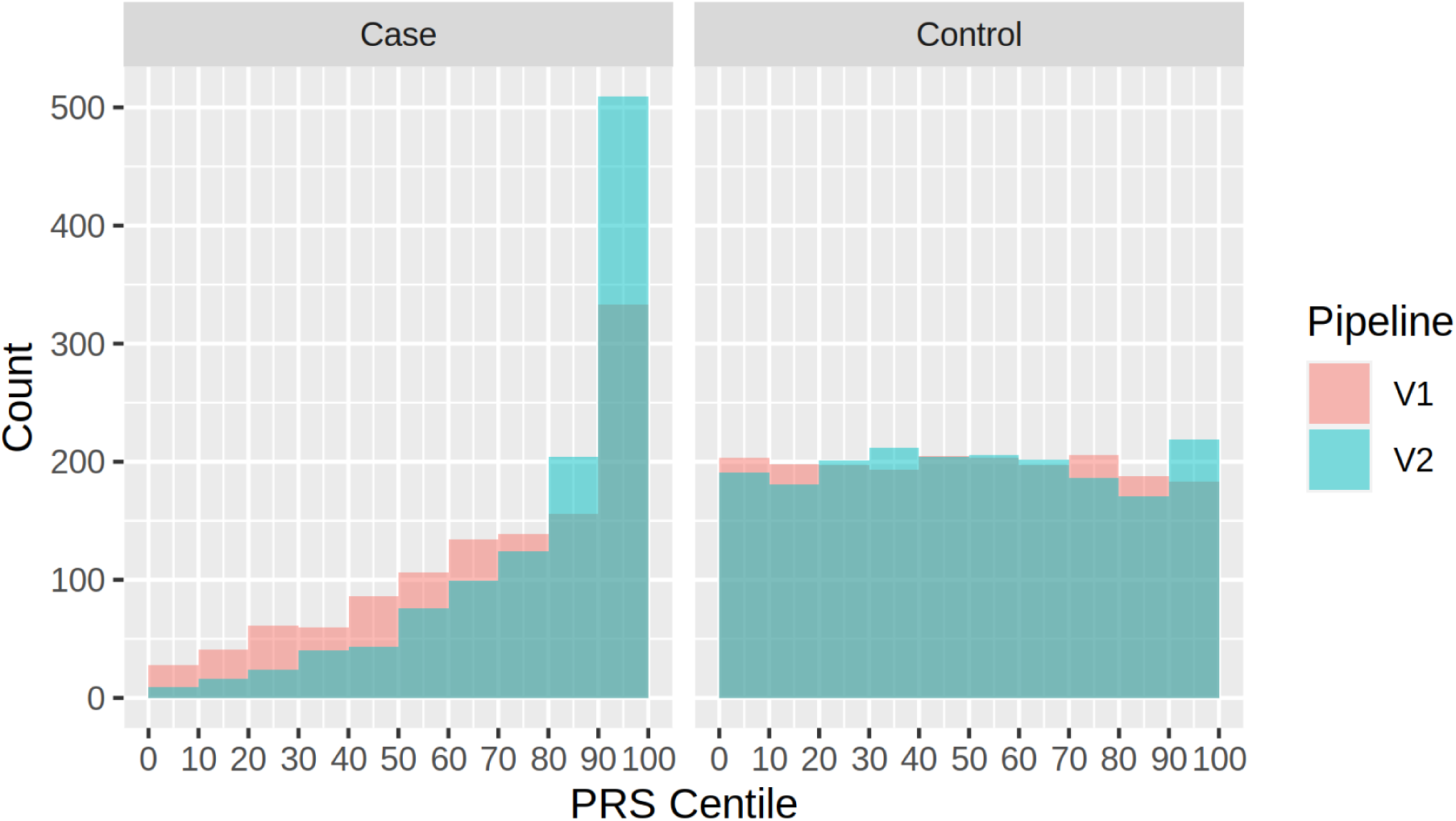
Comparison between old and new PRS in cases and in controls. Comparison between PRS centiles for new PRS (V2) and old PRS (V1) separately in ANZRAG cases and controls.

**Figure S2:**
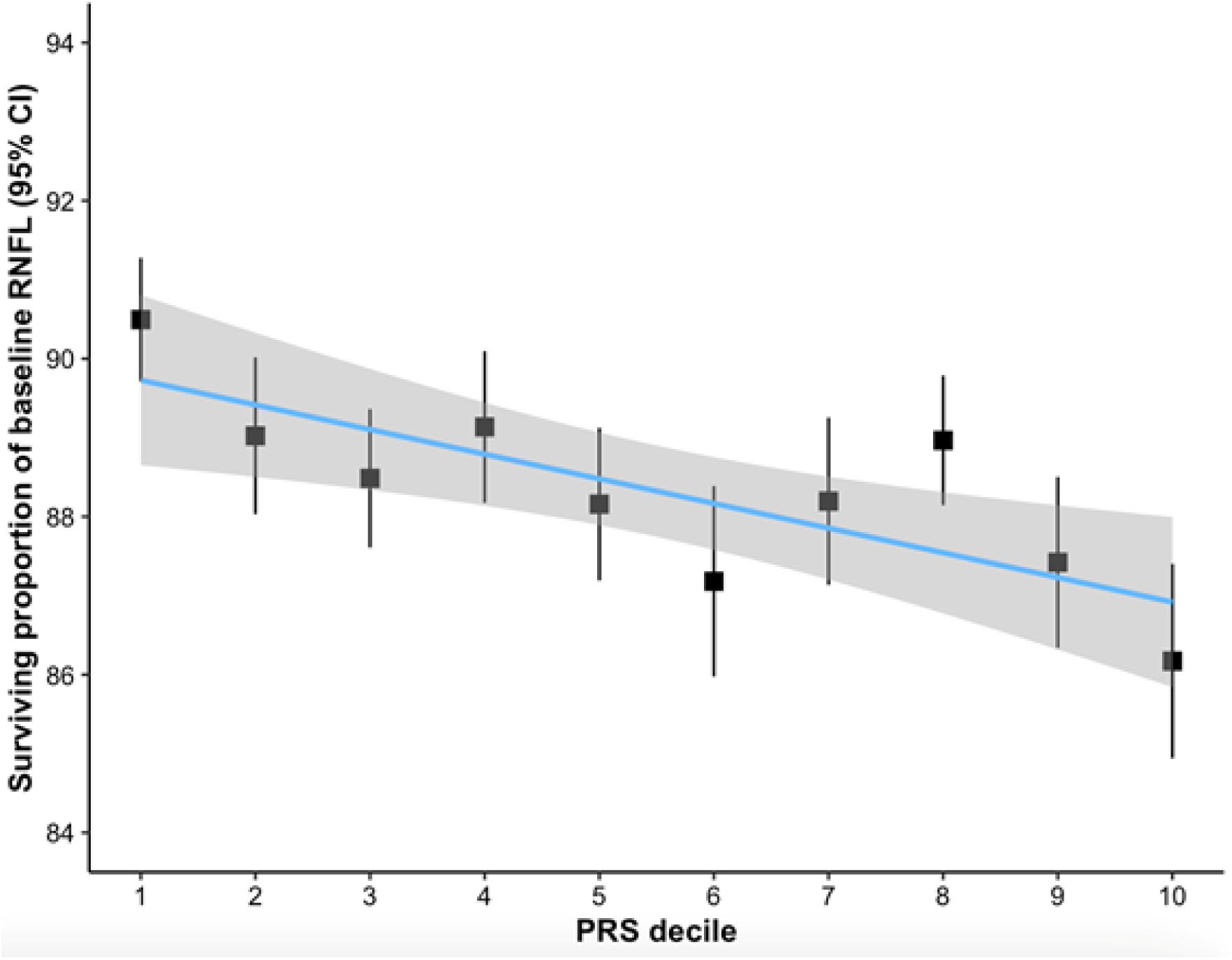
Proportion of preserved baseline retinal nerve fibre layer for PROGRESSA participants using previous PRS (v1) Proportion of preserved baseline retinal nerve fibre layer for PROGRESSA participants with suspect or early manifest glaucoma using previous PRS (v1). The remaining retinal nerve fibre layer proportion is calculated for the most affected quadrant of the most affected eye of each patient — as determined by optical coherence tomography scans at baseline and the latest follow-up scan. The novel PRS predicted the proportion of preserved baseline retinal nerve fibre layer (Figure 3A), with an almost two-fold larger decrease per-decile compared with the previous PRS (shown in the above figure). Each decile change in the new and old PRSs led to −0.52% and −0.3% changes, respectively, in surviving baseline RNFL (difference 0.22%, standard deviation of difference 0.11%, P=0.044).

**Figure S3.**
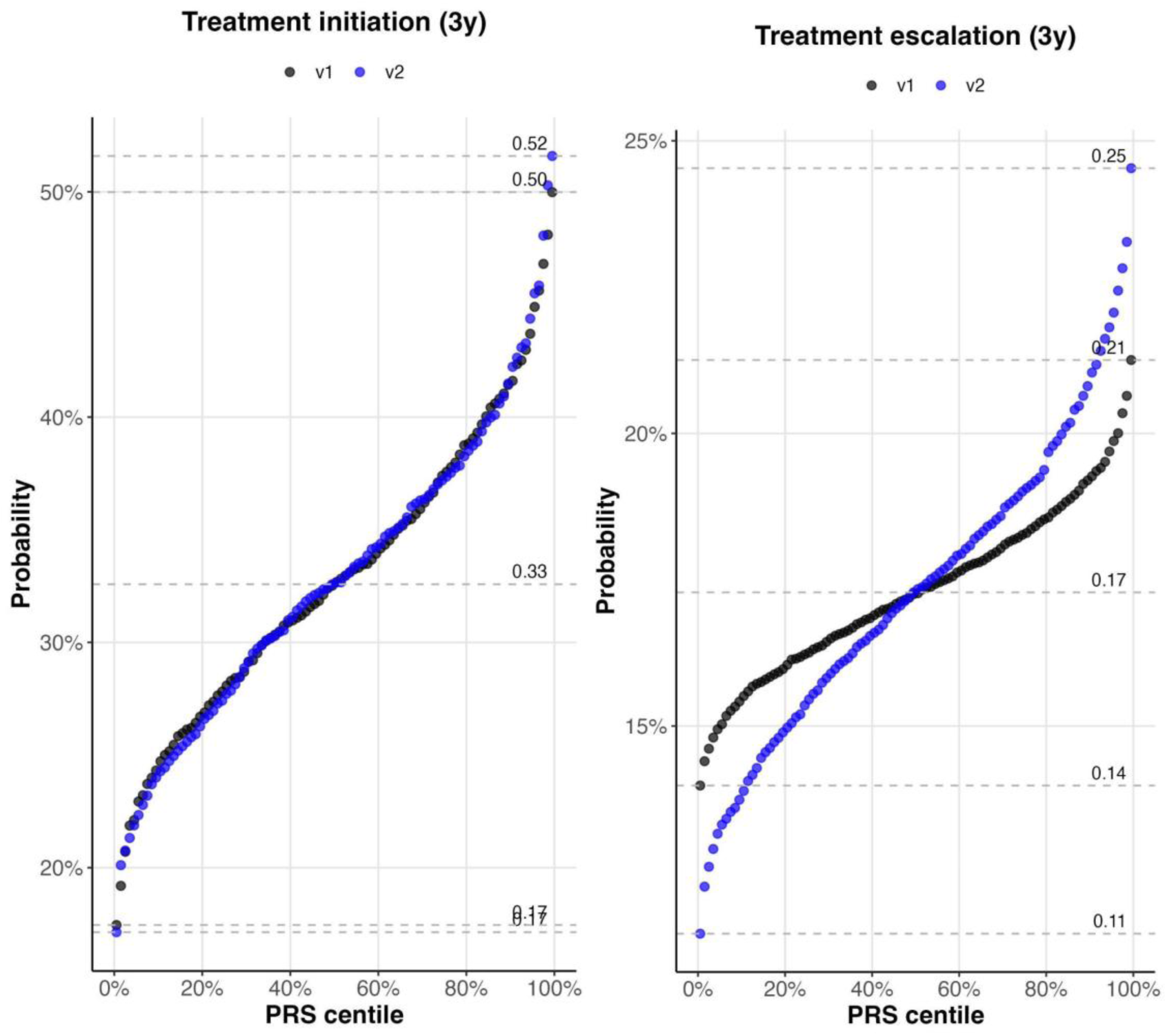
Prediction of treatment initiation or escalation in PROGRESSA. Prediction of treatment initiation or escalation at 3 years in PROGRESSA cohort. V2 is new PRS; V1 is previous PRS.

**Figure S4.**
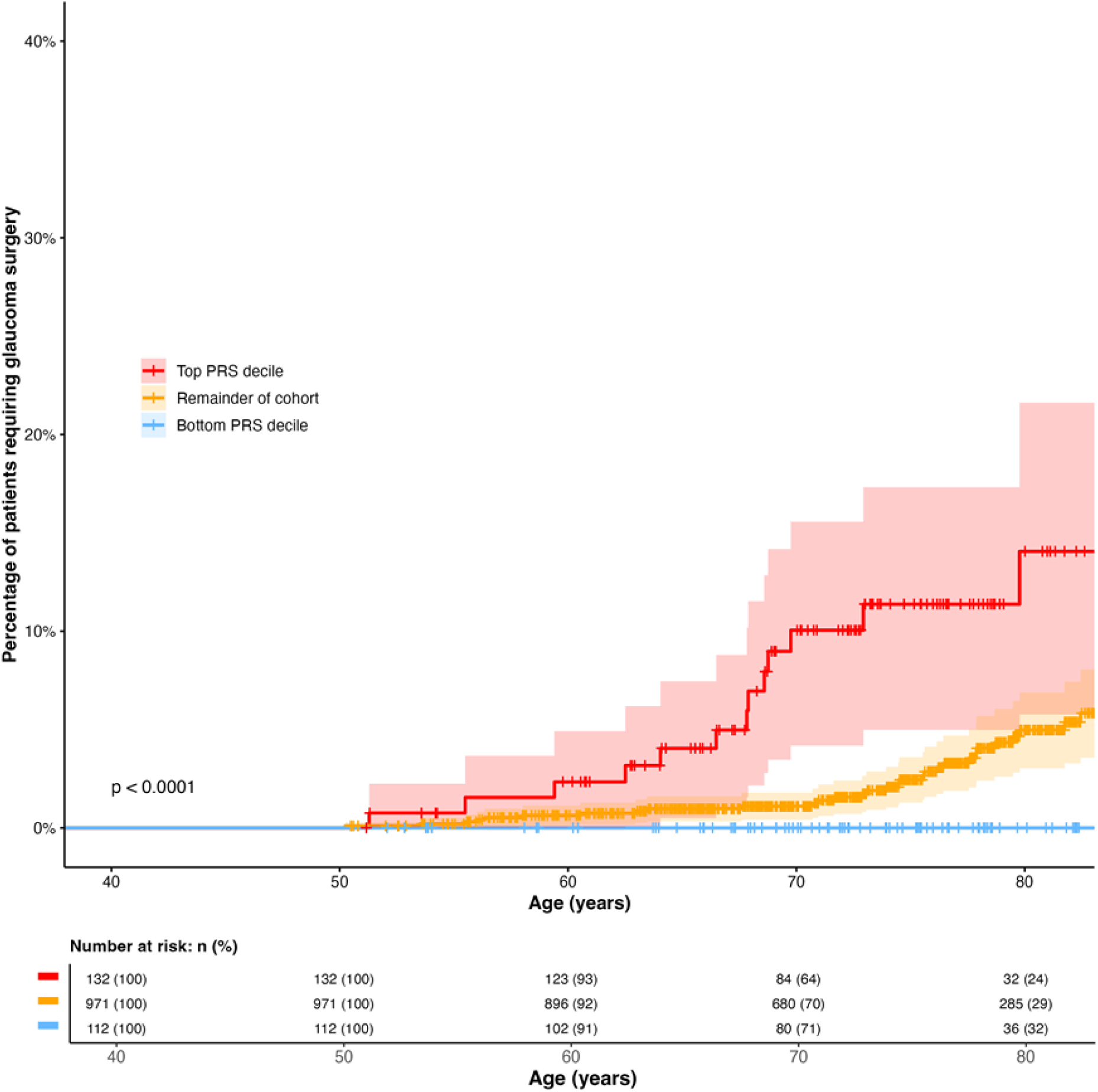
Prediction of incident surgery in prospectively collected PROGRESSA participants. PROGRESSA: Prediction of incident surgery in prospectively collected participants with suspected or early manifest glaucoma using previous PRS (v1). PRS is expressed as a decile internally derived from the PROGRESSA cohort. Red represents participants with a PRS percentile > 90th, yellow represents participants with a PRS percentile 10th-89th, blue represents participants with a PRS percentile < 10th. Solid lines represent the median survival of the group, with 95% CI shaded in the respective color.

**Figure S6.**
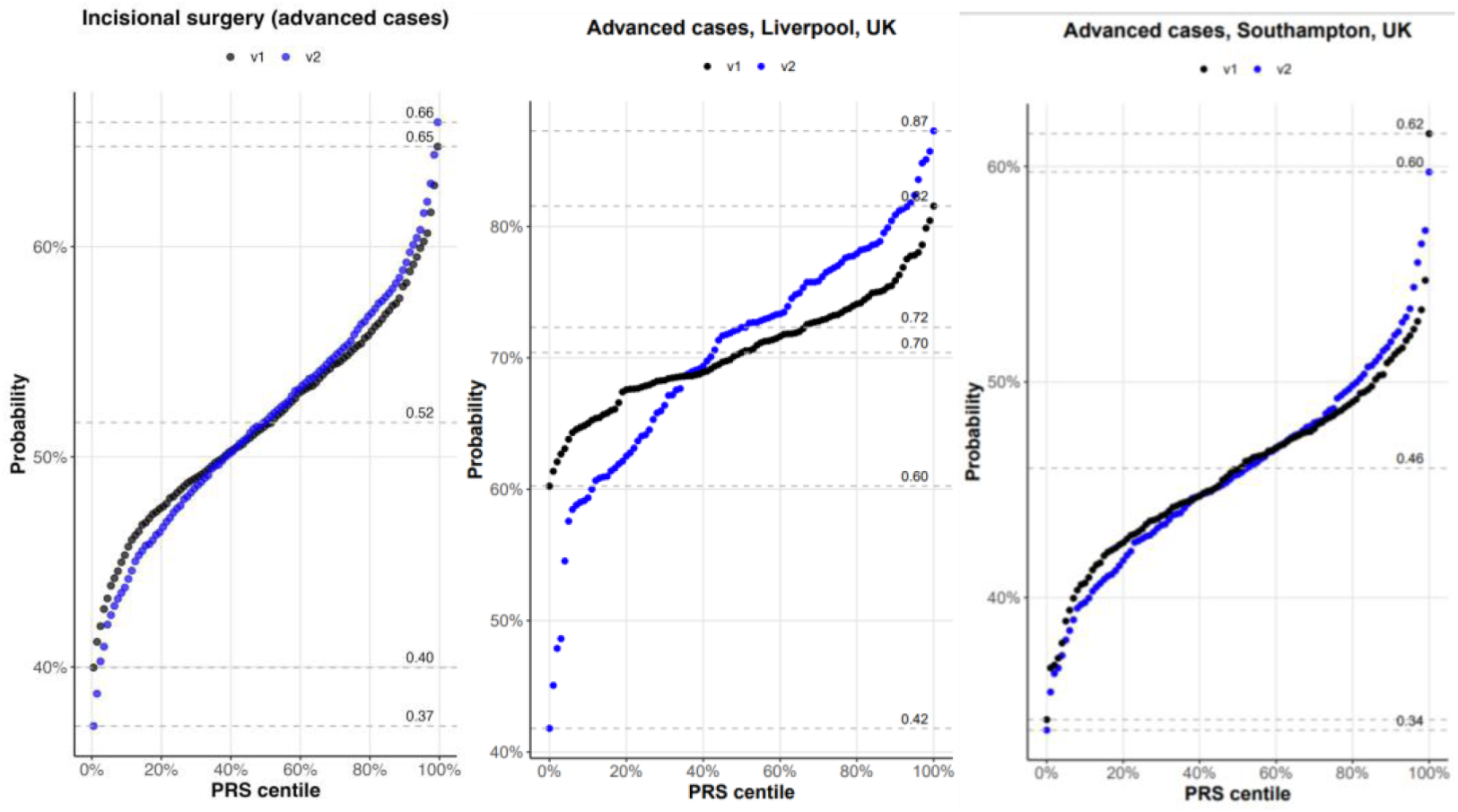
Surgery in advanced glaucoma patients. Results for old PRS (v1); prediction of need for trabeculectomy in advanced glaucoma cases from Australian ANZRAG cohort (N=1434), Liverpool cohort (Willoughby) (N=95) and Southampton (Lotery) cohort (N=299).

**Figure S7:**
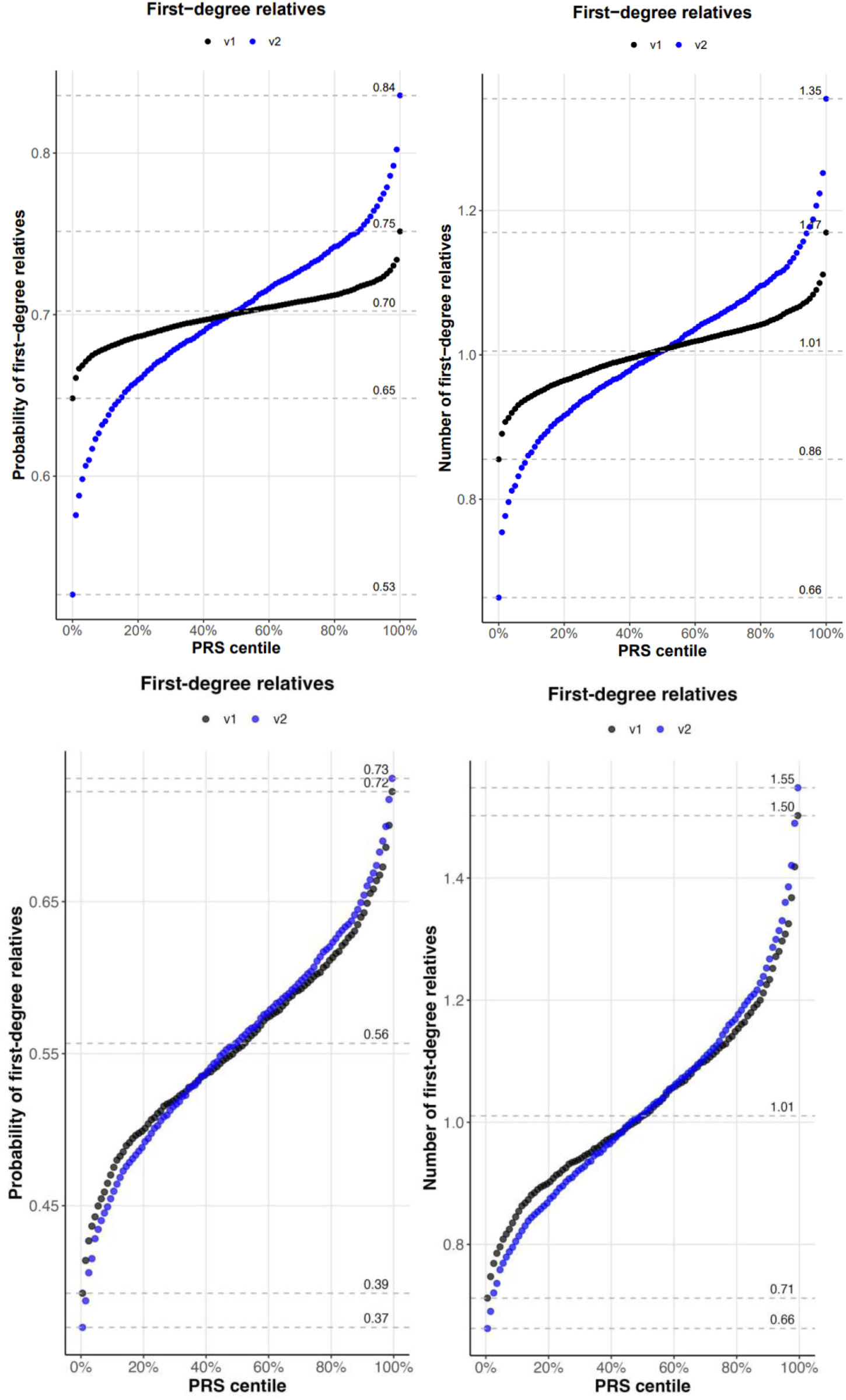
Prediction of probability of having family history. Prediction of probability of having family history (yes/no, number of relatives). Top row: Genetics of Glaucoma Study glaucoma cases. Bottom row: ANZRAG advanced glaucoma cases. In Genetics of Glaucoma, for any first-degree relatives vs not (n=1698) the OR per 1 SD increase in PRS was 1.26 (95% confidence interval 1.14-1.40), P=1.3e-05. The number of first-degree relatives increased by 0.11 (standard error 0.023) per 1 SD increase in PRS, P=1.5e-06. In ANZRAG advanced cases for any first-degree relatives vs not (n=1565) the OR per 1 SD increase in PRS was 1.12 (95% confidence interval 1.08-1.16), P=5.0e-06. The number of first-degree relatives increased by 0.14 (standard error 0.032) per 1 SD increase in PRS, P=1.2e-05.

#### dbGaP Acknowledgement Statement

dbGaP data from accessions phs001312.v1.p1 and phs000238.v1.p1 were used.

The Primary Open-Angle African American Glaucoma Genetics (POAAGG) Study is supported by funding the National Eye Institute, Bethesda, Maryland (grant # 5R01EY023557-03) and the Department of Ophthalmology at the Perelman School of Medicine, University of Pennsylvania, Philadelphia, PA. Funds also come from the

F.M. Kirby Foundation, Research to Prevent Blindness, The Paul and Evanina Bell Mackall Foundation Trust, and the National Eye Institute, National Institutes of Health, Department of Health and Human Services, under eyeGENETM contract Nos. HHSN260220700001C and HHSN263201200001C. The sponsor or funding organization had no role in the design or conduct of this research.

The origin of the dataset is described in the following publication: Charlson ES, Sankar PS, Miller-Ellis E, Regina M, Fertig R, Salinas J, Pistilli M, Salowe RJ, Rhodes AL, Merritt WT, Chua M, Trachtman BT, Gudiseva HV, Collins DW, Chavali VR, Nichols C, Henderer J, Ying GS, Varma R, Jorgenson E, O’Brien JM. (2015) The Primary Open-Angle African-American Glaucoma Genetics (POAAGG) Study: Baseline Demographics. Ophthalmology 122:711-20.

The dataset(s) used for the analyses described in this manuscript were obtained from the NEI Glaucoma Human Genetics Collaboration (NEIGHBOR) Dataset under the dbGaP accession number phs000238. Funding support for NEIGHBOR was provided by the National Eye Institute. We would like to thank NEIGHBOR participants and the NEIGHBOR Research Group for their valuable contribution to this research.

#### Web resources used

GCTA software: http://cnsgenomics.com/software/gcta/

METAL software: http://csg.sph.umich.edu/abecasis/Metal/

Multi-Trait Analysis of GWAS https://github.com/omeed-maghzian/mtag

PLINK software: http://www.cog-genomics.org/plink2

R: https://cran.r-project.org/

UK Biobank: http://www.ukbiobank.ac.uk/

SBayesRC: https://github.com/zhilizheng/SBayesRC

## Abbreviations

AMR: Admixed American/Hispanic
AFR: African
AUC: Area under the curve
ANZRAG: Australian & New Zealand Registry of Advanced Glaucoma
BMES: Blue Mountains Eye Study
CLSA: Canadian Longitudinal Study on Aging
EAS: East Asian
EUR: European
GOG: Genetics of Glaucoma study
GWAS: Genome-wide association study
GBMI: Global Biobank Meta-analysis Initiative
GRADE: Genetic Risk Assessment of Degenerative Eye Disease study
IOP: Intraocular pressure
METAL - software package: Meta-analysis helper
MVP: Million Veteran Program
MTAG - software package: Multiple trait analysis of GWAS
NEI: National Eye Institute Glaucoma Human Genetics Collaboration Heritable Overall Operational Database
OAG: Open-angle glaucoma
PRS: Polygenic risk score
POAAGG: Primary Open-Angle African American Glaucoma Genetics
PROGRESSA: Progression Risk Of Glaucoma: RElevant SNPs with Significant Association study
RNFL: Retinal nerve fibre layer
SNP: Single-nucleotide polymorphisms
SAS: South Asian
UKB: UK Biobank
VCDR: Vertical cup-disc ratio

## REFERENCES

1 Tham Y-C, Li X, Wong TY, Quigley HA, Aung T, Cheng C-Y. Global prevalence of glaucoma and projections of glaucoma burden through 2040: a systematic review and meta-analysis. Ophthalmology 2014; 121: 2081–90.

2 Quigley HA, Broman AT. The number of people with glaucoma worldwide in 2010 and 2020. Br J Ophthalmol 2006; 90: 262–7.

3 Weinreb RN, Khaw PT. Primary open-angle glaucoma. Lancet 2004; 363: 1711–20.

4 Siggs OM, Qassim A, Han X, et al. Association of High Polygenic Risk With Visual Field Worsening Despite Treatment in Early Primary Open-Angle Glaucoma. JAMA Ophthalmol 2022; 141: 73–7.

5 Marshall HN, Mullany S, Han X, et al. High Polygenic Risk Is Associated with Earlier Initiation and Escalation of Treatment in Early Primary Open-Angle Glaucoma. Ophthalmology 2023; 130: 830–6.

6 Fraser S, Bunce C, Wormald R. Risk factors for late presentation in chronic glaucoma. Invest Ophthalmol Vis Sci 1999; 40: 2251–7.

7 Burr JM, Mowatt G, Hernández R, et al. The clinical effectiveness and cost-effectiveness of screening for open angle glaucoma: a systematic review and economic evaluation. Health Technol Assess 2007; 11: iii – iv, ix – x, 1–190.

8 Chou R, Selph S, Blazina I, et al. Screening for Glaucoma in Adults: Updated Evidence Report and Systematic Review for the US Preventive Services Task Force. JAMA 2022; 327: 1998–2012.

9 Chan MPY, Broadway DC, Khawaja AP, et al. Glaucoma and intraocular pressure in EPIC-Norfolk Eye Study: cross sectional study. BMJ 2017; 358: j3889.

10 Dielemans I, Vingerling JR, Wolfs RC, Hofman A, Grobbee DE, de Jong PT. The prevalence of primary open-angle glaucoma in a population-based study in The Netherlands. The Rotterdam Study. Ophthalmology 1994; 101: 1851–5.

11 Zhao J, Solano MM, Oldenburg CE, et al. Prevalence of Normal-Tension Glaucoma in the Chinese Population: A Systematic Review and Meta-Analysis. Am J Ophthalmol 2019; 199: 101–10.

12 Wolfs RC, Klaver CC, Ramrattan RS, van Duijn CM, Hofman A, de Jong PT. Genetic risk of primary open-angle glaucoma. Population-based familial aggregation study. Archives of ophthalmology (Chicago, Ill : 1960) 1998; 116. DOI:10.1001/archopht.116.12.1640.

13 Craig JE, Han X, Qassim A, et al. Multitrait analysis of glaucoma identifies new risk loci and enables polygenic prediction of disease susceptibility and progression. Nat Genet 2020; 52. DOI:10.1038/s41588-019-0556-y.

14 Hao L, Kraft P, Berriz GF, et al. Development of a clinical polygenic risk score assay and reporting workflow. Nat Med 2022; 28: 1006–13.

15 Lennon NJ, Kottyan LC, Kachulis C, et al. Selection, optimization and validation of ten chronic disease polygenic risk scores for clinical implementation in diverse US populations. Nat Med 2024; 30: 480–7.

16 Patel AP, Wang M, Ruan Y, et al. A multi-ancestry polygenic risk score improves risk prediction for coronary artery disease. Nat Med 2023; 29: 1793–803.

17 Wang K, Gaitsch H, Poon H, Cox NJ, Rzhetsky A. Classification of common human diseases derived from shared genetic and environmental determinants. Nat Genet 2017; 49: 1319–25.

18 Sanfilippo PG, Hewitt AW, Hammond CJ, Mackey DA. The heritability of ocular traits. Surv Ophthalmol 2010; 55: 561–83.

19 Choquet H, Paylakhi S, Kneeland SC, et al. A multiethnic genome-wide association study of primary open-angle glaucoma identifies novel risk loci. Nat Commun 2018; 9: 2278.

20 Gharahkhani P, Jorgenson E, Hysi P, et al. Genome-wide meta-analysis identifies 127 open-angle glaucoma loci with consistent effect across ancestries. Nat Commun 2021; 12: 1258.

21 Gharahkhani P, Burdon KP, Fogarty R, et al. Common variants near ABCA1, AFAP1 and GMDS confer risk of primary open-angle glaucoma. Nat Genet 2014; 46: 1120–5.

22 Han X, Hewitt AW, MacGregor S. Predicting the Future of Genetic Risk Profiling of Glaucoma: A Narrative Review. JAMA Ophthalmol 2021; 139: 224–31.

23 Leske MC, Heijl A, Hyman L, Bengtsson B, Komaroff E. Factors for progression and glaucoma treatment: the Early Manifest Glaucoma Trial. Curr Opin Ophthalmol 2004; 15: 102–6.

24 Garway-Heath DF, Crabb DP, Bunce C, et al. Latanoprost for open-angle glaucoma (UKGTS): a randomised, multicentre, placebo-controlled trial. Lancet 2015; 385: 1295–304.

25 Mansouri K, Tanna AP, De Moraes CG, Camp AS, Weinreb RN. Review of the measurement and management of 24-hour intraocular pressure in patients with glaucoma. Surv Ophthalmol 2020; 65: 171–86.

26 Turley P, Walters RK, Maghzian O, et al. Multi-trait analysis of genome-wide association summary statistics using MTAG. Nat Genet 2018; 50: 229–37.

27 Zheng Z, Liu S, Sidorenko J, et al. Leveraging functional genomic annotations and genome coverage to improve polygenic prediction of complex traits within and between ancestries. Nat Genet 2024; 56: 767–77.

28 Willer CJ, Li Y, Abecasis GR. METAL: fast and efficient meta-analysis of genomewide association scans. Bioinformatics 2010; 26: 2190–1.

29 R Core Team. R: A Language and Environment for Statistical Computing. 2017. https://www.R-project.org/.

30 Purcell S, Neale B, Todd-Brown K, et al. PLINK: a tool set for whole-genome association and population-based linkage analyses. Am J Hum Genet 2007; 81: 559–75.

31 Robin X, Turck N, Hainard A, et al. pROC: an open-source package for R and S+ to analyze and compare ROC curves. BMC Bioinformatics. 2011; 12: 77.

32 Liu Q, Davis J, Han X, et al. Cost-effectiveness of polygenic risk profiling for primary open-angle glaucoma in the United Kingdom and Australia. Eye (Lond) 2023; 37: 2335–43.

33 Han X, Steven K, Qassim A, et al. Automated AI labeling of optic nerve head enables insights into cross-ancestry glaucoma risk and genetic discovery in >280,000 images from UKB and CLSA. Am J Hum Genet 2021; 108: 1204–16.

34 Wand H, Kalia SS, Helm BM, et al. Clinical genetic counseling and translation considerations for polygenic scores in personalized risk assessments: A Practice Resource from the National Society of Genetic Counselors. J Genet Couns 2023; 32: 558–75.

35 Young M-A, Yanes T, Cust AE, et al. Human Genetics Society of Australasia Position Statement: Use of Polygenic Scores in Clinical Practice and Population Health. Twin Res Hum Genet 2023; 26: 40–8.

36 Mitchell P, Smith W, Attebo K, Healey PR. Prevalence of open-angle glaucoma in Australia. The Blue Mountains Eye Study. Ophthalmology 1996; 103: 1661–9.

37 Shaikh Y, Yu F, Coleman AL. Burden of undetected and untreated glaucoma in the United States. Am J Ophthalmol 2014; 158: 1121–9.e1.

## Supplementary References

1 Zhang H, Zhan J, Jin J, et al. A new method for multiancestry polygenic prediction improves performance across diverse populations. Nat Genet 2023; 55: 1757–68.

2 Verma A, Huffman JE, Rodriguez A, et al. Diversity and scale: Genetic architecture of 2068 traits in the VA Million Veteran Program. Science 2024; 385: eadj1182.

3 Lo Faro V, Bhattacharya A, Zhou W, et al. Novel ancestry-specific primary open-angle glaucoma loci and shared biology with vascular mechanisms and cell proliferation. Cell Rep Med 2024; 5: 101430.

4 Han X, Gharahkhani P, Hamel AR, et al. Large-scale multitrait genome-wide association analyses identify hundreds of glaucoma risk loci. Nat Genet 2023; 55: 1116–25.

5 Bycroft C, Freeman C, Petkova D, et al. Genome-wide genetic data on ∼500,000 UK Biobank participants. 2017. DOI:10.1101/166298.

6 Bonnemaijer PWM, Leeuwen EMV, Iglesias AI, et al. Multi-trait genome-wide association study identifies new loci associated with optic disc parameters. Communications biology 2019; 2. DOI:10.1038/s42003-019-0634-9.

7 Han X, Steven K, Qassim A, et al. Automated AI labeling of optic nerve head enables insights into cross-ancestry glaucoma risk and genetic discovery in >280,000 images from UKB and CLSA. Am J Hum Genet 2021; 108: 1204–16.

8 Gharahkhani P, Burdon KP, Fogarty R, et al. Common variants near ABCA1, AFAP1 and GMDS confer risk of primary open-angle glaucoma. Nat Genet 2014; 46: 1120–5.

9 Mitchell P, Smith W, Attebo K, Healey PR. Prevalence of open-angle glaucoma in Australia. The Blue Mountains Eye Study. Ophthalmology 1996; 103: 1661–9.

10 Gharahkhani P, He W, Diaz Torres S, et al. Study profile: the Genetics of Glaucoma Study. BMJ Open 2023; 13: e068811.

11 Hollitt GL, Qassim A, Thomson D, et al. Genetic Risk Assessment of Degenerative Eye Disease (GRADE): study protocol of a prospective assessment of polygenic risk scores to predict diagnosis of glaucoma and age-related macular degeneration. BMC Ophthalmol 2023; 23: 431.

12 Olsen CM, Green AC, Neale RE, et al. Cohort profile: the QSkin Sun and Health Study. Int J Epidemiol 2012; 41: 929–929i.

14 Gharahkhani P, Jorgenson E, Hysi P, et al. Genome-wide meta-analysis identifies 127 open-angle glaucoma loci with consistent effect across ancestries. Nat Commun 2021; 12: 1258.

